# IBDex: A Frozen Patient-Scale Atlas for Bulk Transcriptomics in Inflammatory Bowel Disease

**DOI:** 10.64898/2026.10.20.26365993

**Authors:** Ajith Kumar Madeswaran, Animesh Acharjee

## Abstract

**Background & Aims:** Bulk mucosal RNA sequencing in inflammatory bowel disease (IBD) is analyzed one cohort at a time, so each study rebuilds its own molecular scale. We aimed to build a frozen atlas that new cohorts could use without retraining.

**Methods:** We built IBDex from 3,168 intestinal biopsies from 1,680 patients with Crohn’s disease, ulcerative colitis or no IBD, drawn from four cohorts. Patients were kept within a single training, validation or test partition. A tissue-aware conditional variational autoencoder with adversarial suppression of cohort identity was trained on a 3,000-gene interface. The scaling, encoder, decoder, pathway basis and phenotype directions were then frozen. Phenotype-anchored counterfactual perturbation (PACP) produced anatomy-conditioned gene-response maps. We tested the atlas without parameter updates in three unseen IBD cohorts, a longitudinal olamkicept trial, and cohorts with primary sclerosing cholangitis (PSC) or colorectal cancer (CRC).

**Results:** In held-out data, dataset identity fell to near chance (balanced accuracy 0.307 vs 0.250). Anatomy, diagnosis and inflammation remained recoverable (inflammation AUC 0.82). Marginal differential expression conflated inflammation and disease state (ρ = 0.834). PACP separated them (ρ = −0.161), with inflammation driven by TNF–NF-κB, and disease state by proliferative, type I interferon and metabolic programs. In 93 patients in combined endoscopic and histological remission, inflammation approached controls (AUC 0.58), but disease state stayed displaced (AUC 0.725, P = 1.8 × 10□^1^□). The frozen atlas recovered clinical, single-cell-defined and disease-behavior structure in unseen cohorts. It also tracked responder trajectories during olamkicept therapy and separated PSC (patient-level AUC 0.794) and CRC (AUC 0.938– 1.000). Where de novo differential expression became unreliable, transferred response maps stayed reproducible (ρ ≥ 0.89).

**Conclusions:** IBDex turns historical bulk biopsy data into a reusable reference. It shows that deep remission moves inflammation toward control levels but leaves a persistent molecular disease state.

## Introduction

Intestinal tissue is among the most extensively profiled human tissues, yet bulk RNA sequencing is analyzed one cohort at a time. Public repositories contain thousands of mucosal biopsies spanning cross-sectional, longitudinal and interventional studies [1–3], but each study rebuilds normalization, technical correction, representation, differential contrasts and pathway interpretation. The resulting molecular scale belongs to the cohort rather than to the tissue. Historical data accumulate without becoming a reference.

This limitation is especially consequential in inflammatory bowel disease (IBD), where therapeutic options have expanded faster than the ability to determine which mechanism is active in an individual mucosa. Non-response remains common across biologic therapies [4]. Selecting Therapeutic Targets in Inflammatory Bowel Disease II (STRIDE-II) sharpened treatment goals toward remission, biomarker normalization and endoscopic healing [5], yet these endpoints primarily measure inflammatory resolution. They do not establish whether deep remission restores non-IBD mucosal biology or leaves a persistent molecular disease state.

Mucosal transcriptomics has repeatedly identified mechanisms linked to disease activity and treatment resistance. Oncostatin M marks a stromal-myeloid program associated with anti-TNF resistance [6], and single-cell profiling resolves treatment-resistant cellular modules [7]. The challenge is no longer detecting signal, but measuring the same biology on a scale that persists across studies. Current signals remain defined on study-specific scales and must be reconstructed before they can be measured elsewhere.

Single-cell genomics addressed the analogous infrastructure problem by separating atlas construction from atlas use. The Human Cell Atlas established persistent molecular maps [8], scVI enabled scalable latent references [9], scArches formalized query mapping [10], and scGen showed that latent perturbation vectors transfer across contexts [11]. In IBD, scIBD demonstrated that published single-cell studies can form a disease-focused meta-atlas [12]. Molecular atlases can therefore outlive their construction datasets.

Bulk tissue requires a different atlas because it preserves patient-level variation while mixing anatomy, disease state and changing cellular composition. What remains missing is a reusable bulk-tissue reference with a fixed gene interface, reference scaling, pathway vocabulary, validated phenotype directions and calibrated distributions that allow a biopsy to be interpreted without rebuilding the system. Conditional generative models provide the necessary machinery [13], but bulk transcriptomics still lacks a reference frame that persists beyond its construction cohort.

We therefore developed IBDex as a frozen patient-scale tissue atlas. The atlas was built with a tissue-aware conditional variational autoencoder, and the completed gene interface, scaling, encoder, decoder, pathway basis and phenotype directions were frozen together before application to subsequent cohorts. Anatomy remains an explicit condition rather than nuisance variation, allowing the same molecular displacement to be queried in colon and ileum. Future cohorts are projected rather than rebuilt and cannot alter the atlas. Using this fixed reference, we asked whether inflammation could be separated from a persistent disease state, whether deep remission normalized both processes, and whether the same molecular scale transferred across unseen cohorts, diseases and an interventional trial. The contribution is not another embedding. It is a frozen patient-scale tissue atlas that converts historical bulk transcriptomes into a reusable molecular reference.

## Materials and Methods

### Cohort assembly and patient-blocked partitioning

IBDex was constructed from four adult human intestinal bulk RNA-sequencing resources: GSE193677 [1], IBDome [3], GSE165512 and the Integrative Human Microbiome Project (iHMP2) [2]. Eligible samples were adult mucosal biopsies with Crohn’s disease, ulcerative colitis or non-IBD control status, a recorded anatomical site and patient linkage. Metadata were harmonized to seven sites: rectum, left colon, right colon, transverse colon, sigmoid colon, colon unspecified and ileum, and to colon or ileum. The reference comprised 3,168 biopsies from 1,680 patients: 1,510 Crohn’s disease, 1,073 ulcerative colitis and 585 controls, including 2,198 colonic and 970 ileal samples.

Genes were mapped to HGNC symbols and transformed within cohort to log_2_(CPM + 1), leaving 21,912 genes shared by all cohorts. Patients were partitioned with stratification by cohort, diagnosis and compartment, and all biopsies from one patient remained together. Training contained 2,222 samples from 1,175 patients, validation 479 from 253 and held-out testing 467 from 252 (Supplementary Table 1).

Inputs were standardized with training-partition statistics (equation (1)),

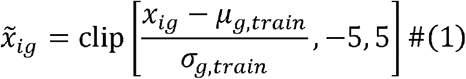

where μ_g,train_ and σ_g,train_ are the training mean and standard deviation. The same quantities and a fixed ±5 s.d. clipping rule were applied to validation, test and external samples, which did not contribute to scaling.

Gene selection and scaling used training data only. Validation selected the checkpoint and then joined training data for final decoder fitting. The test partition remained excluded from these steps and was used for representation probes and phenotype validation. After model freezing, all 3,168 reference biopsies were used to construct and calibrate the pathway coordinates.

### Frozen gene interface and baseline comparison

With the corpus fixed, gene selection defined the measurement interface. A training-only compatibility audit classified the 21,912 shared genes by cross-cohort behavior and presence– absence artifacts (Supplementary Table 4). Prespecified expression, detection, reproducibility, within-tissue cohort-confounding, presence–absence and residual-estimability gates, followed by a declared low-mean rescue, yielded 9,084 eligible genes. Sex-linked, mitochondrial and hemoglobin genes were excluded and immunoglobulin variable regions constrained (Supplementary Tables 5–8).

Eligible genes were ranked by the prespecified additive score in equation (2):

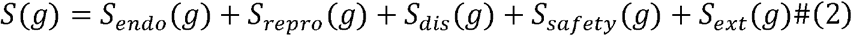

The score combined residual biological heterogeneity, cross-cohort reproducibility, disease association, dataset safety and capped external evidence, with maximum contributions of 40, 20, 15, 10 and 15 points (Supplementary Tables 6 and 7).

External evidence was defined by equation (3):

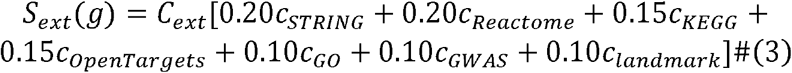

The fixed evidence sources were STRING, Reactome [14], KEGG [15], Open Targets, Gene Ontology Biological Process, the NHGRI-EBI GWAS Catalog and a curated landmark indicator. Absent annotations contributed zero (Supplementary Table 7).

All 213 mechanism and therapeutic landmark genes were audited without changing eligibility or rank. Of 53 absent from the final panel, 47 failed batch- or tissue-safety gates and six failed expression criteria. None was force-added (Supplementary Table 8). The 3,000 highest-scoring genes formed the fixed interface and were not reselected for any downstream analysis.

Two conventional representations were constructed on the same frozen 3,000-gene interface before generative modeling. Baseline 1, principal-component analysis (PCA), was fitted to the standardized expression matrix and the leading 50 components retained. For visualization, these components were embedded with Uniform Manifold Approximation and Projection (UMAP) using 15 neighbors and minimum distance 0.30. The second applied per-dataset location-and-scale harmonization with identical UMAP settings [16]. Because the matrix was log_2_(CPM + 1), this comparator is termed ComBat-seq-style rather than a refit of the original count-level negative-binomial model. Both baselines were evaluated on the matrix used for IBDex.

### Adversarial tissue-aware CVAE

The fixed interface was then used to train a tissue-aware conditional variational autoencoder (CVAE). For standardized expression x ∈ □^3000^, the encoder condition c_e_ ∈ {0,1}^6^ contained broad tissue and four dataset indicators. The decoder condition c_d_ ∈ {0,1}^11^ contained the four dataset indicators and seven anatomical sites. The posterior and decoder were defined by equations (4) and (5):

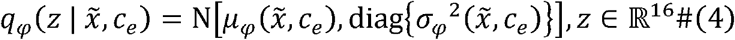

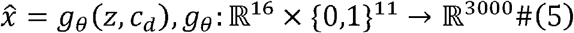

Encoder and decoder each used two 512-unit layers with layer normalization, GELU activation and dropout 0.10. Training used reparameterized latent sampling (equation (6)),

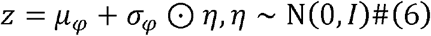

whereas downstream analyses used the deterministic posterior mean μ_φ_.

Dataset identity was masked from the encoder with probability 0.3 per sample, but retained by the decoder together with full anatomical site. This asymmetric conditioning required a provenance-independent latent code while preserving anatomy as an explicit decoder variable. Three gradient-reversal heads [17] suppressed residual provenance: nonlinear and linear four-class dataset classifiers and a seven-class anatomical-site classifier. For strength γ, the forward operation was

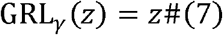

and its back-propagated derivative was

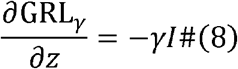

The dataset heads constrained nonlinear and linear provenance, while the site head discouraged fine anatomical identity in the latent code without removing anatomy from the decoder.

For mini-batch size B, reconstruction error was the sample-averaged squared error across 3,000 genes (equation (9)),

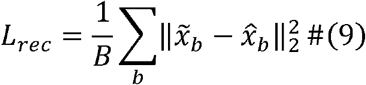

and the variational penalty was the diagonal-Gaussian Kullback–Leibler divergence from a standard normal prior (equation (10)),

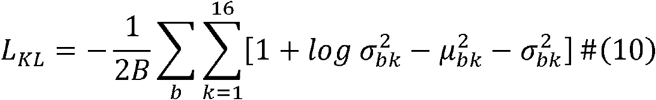

giving the complete objective in equation (11):

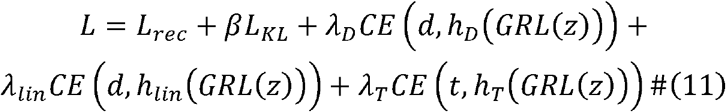

The reported configuration used β_max_ = 1, λ_D_ = 120, λ_lin_ = 120 and λ_T_ = 80. These terms and the corresponding reversal strengths were linearly ramped from epochs 1 to 120.

Batches contained 32 samples balanced across cohorts, with cohort and tissue sampling weights of 1.0 and 0.25. AdamW used learning rate 6 × 10^−4^, weight decay 3.1 × 10^−4^ and gradient clipping at 5.0. Training ran for at most 300 epochs with patience 30 and no eligible checkpoint before epoch 150.

A development-stage hyperparameter search selected the configuration by validation loss (Supplementary Table 9). At epoch 179, encoder and decoder parameters, the gene panel, training statistics and clipping rule were fixed and no future analysis updated them.

### Latent-space validation

After the model was frozen, post-training information probes quantified the information retained in the latent space without influencing model selection. Balanced multinomial logistic-regression probes were fitted to standardized training latents and evaluated in validation and test data. Targets were dataset, seven-level site, colon versus ileum, diagnosis and inflammatory activity. Confidence intervals used 1,999 patient-level bootstrap resamples and significance 1,999 patient-level label permutations, with Benjamini–Hochberg correction [18] within the probe family. Per-dimension variance partitioning quantified dataset partial R^2^ after adjustment for diagnosis and anatomical site.

Consensus clustering [19] tested for stable discrete structure rather than imposing endotypes. Prespecified k-means, Gaussian-mixture, spectral and Leiden [20] solutions were generated for k = 2,…,8 in colon, ileum and the mixed reference (Supplementary Table 10). For run h, M_h_(i,j) indicated co-membership and I_h_(i,j) joint inclusion. Consensus similarity was

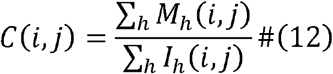

Hierarchical clustering of C produced candidate regions, evaluated by silhouette, Calinski– Harabasz and Davies–Bouldin indices under prespecified size constraints. A 250-permutation null and bootstrap Jaccard and adjusted Rand indices assessed stability.

### Decoder-derived pathway atlas

We next converted the local geometry of the frozen decoder into pathway coordinates. For anatomical condition t, the mean Jacobian J_t_ ∈ □^3000×16^ was estimated across tissue-matched reference biopsies. Column j used centered finite differences with ε = 10^−2^ (equation (13)),

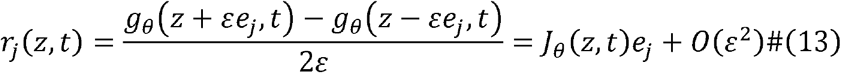

For each Hallmark [21], Reactome [14] or KEGG [15] pathway, genes were ranked by each response vector and tested by preranked GSEA [22]. Sets with 10–500 panel genes were retained. Significance used 500 permutations with collection-specific FDR control [18]. The resulting enrichment scores defined the tissue-specific direction

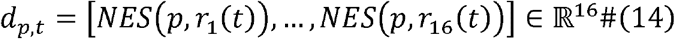

and biopsy i was projected onto its tissue-specific direction and calibrated against the complete 3,168-biopsy reference (equation (15)),

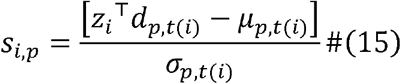

where μ_p,t_ and σ_p,t_ are the tissue-specific reference mean and standard deviation. Thus, training data selected and scaled the interface, the test partition did not fit the decoder, and the complete reference subsequently defined Jacobians, pathway directions and calibration distributions. A prespecified panel of 16 coordinates covered inflammatory signaling, fibrosis and remodeling, growth and stress, bile-acid biology and barrier glycosylation (Supplementary Tables 24 and 25). Coordinates were corroborated against full-transcriptome and panel-matched ssGSEA [23] across all reference biopsies by compartment-specific Spearman correlation. Fixed reliability bands were applied without using concordance to select coordinates.

### Phenotype axes and PACP

To connect the pathway atlas to clinical phenotypes, prespecified contrasts were converted into axes in the frozen latent reference. For groups A and B, the initial unit direction was

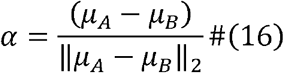

Fourteen candidates with at least 25 biopsies per group were orthogonalized in a fixed priority order, yielding seven directions above the residual-norm threshold in equation (17) (Supplementary Tables 14 and 15).

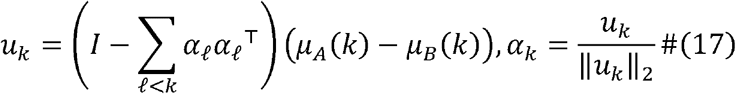

Candidates were retained when residual norm exceeded 0.20. Directions were evaluated without refitting in patient-disjoint validation and test data. Signs were fixed by training contrasts, and five of seven directions passed held-out permutation validation. Inflammation and active-versus-inactive contrasts used distinct annotations. Disease state compared Crohn’s disease and ulcerative colitis jointly with controls.

Phenotype-anchored counterfactual perturbation (PACP) measured the decoder’s local gene-level response along a validated unit phenotype direction α. For anatomical condition t and evaluation states Z,

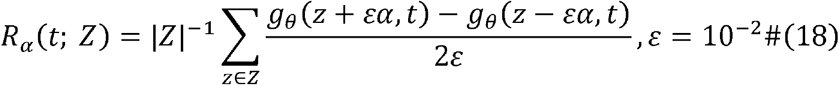

R (t;Z) ∈ □^3000^ is a signed response map, not a reconstructed profile. Site-level Jacobians used rectum (n = 961), right colon (n = 498), left colon (n = 292), sigmoid colon (n = 203), transverse colon (n = 113), colon unspecified (n = 131) and ileum (n = 970). Compartment operators used count-weighted site aggregation. Robustness was checked across ε = 10^−3^ to 10^−1^, equal site weights and elementwise medians.

### Response-map benchmarking

The resulting response maps were benchmarked first at pathway level. Signed PACP loadings for inflammation and disease state were tested in colon and ileum by weighted signed-rank enrichment. The primary p = 1 analysis and p = 0 sensitivity analysis included 44 Hallmark sets with at least 10 panel genes. Significance used 2,000 same-size random sets with Benjamini– Hochberg correction within phenotype-by-compartment families (Supplementary Table 26).

Response maps were compared by signed Spearman correlation, directional agreement and top-k same-sign overlap after ranking absolute magnitude. A value of k = 100 was prespecified for PACP versus differential expression. Colon and ileum PACP operators were estimated from the same evaluation states under their respective anatomical conditions. All 3,000 genes were compared by signed rank and direction agreement. Reversals were stratified by the better response-magnitude rank to distinguish strong-response changes from reversals in the weak tail. Differential expression (DE) used the same frozen panel, patients and contrasts as PACP. Mann– Whitney U was converted to AUC − 0.5, pooled and within tissue. PACP and DE were compared by signed Spearman ρ, direction agreement and top-100 same-sign overlap.

Patient-blocked split halves assessed reproducibility. DE was re-estimated in each half, whereas PACP remained frozen and only evaluation states changed. Thirty internal splits were used for the primary comparison, with denser DE resampling where required. Per-gene PACP stability used 200 patient-level bootstrap draws within tissue. Two-sided sign-flip P values were Benjamini–Hochberg corrected within phenotype-by-tissue families [18].

### Clinical and external validation

Clinical validation first tested remission using the locked inflammation and disease-state directions without retraining or recalibration. Concordant endoscopic and histological remission was compared with concordantly active disease and controls after reducing repeated biopsies to patient medians. The principal analysis included 93 remission, 252 active and 241 control patients. Pairwise differences used two-sided Mann–Whitney U, AUC-based direction and patient-level bootstrap intervals. Endoscopic-only and histological-only definitions were sensitivity analyses.

External datasets were mapped to the 3,000-gene interface, standardized with training μ_g,train_ and σ_g,train_ and clipped to [−5,5]. Missing genes were assigned the standardized training mean. Dataset indicators were set to zero and deterministic posterior means supplied latent coordinates. No interface, scaling, encoder, decoder, pathway basis or calibration quantity was updated. Three unseen IBD cohorts comprised 115 biopsies. In GSE130038 [24], all 16 coordinates were screened against disease duration in 26 patients, and the top coordinate was independently checked by full-transcriptome ssGSEA.

In GSE235236 [25], five cell-state programs defined from the source study before comparison with IBDex were scored in 56 bulk biopsies. The signatures comprised M2 and IDA macrophage programs (the top 10 genes by mean expression difference for each), the reported nine-gene inflammatory-fibroblast panel, the top cluster-specific N3 interferon-neutrophil genes and four reported pan-neutrophil markers (Supplementary Table 22). All five programs were scored across the complete matrix using rank-normalized ssGSEA with minimum gene-set size 3, and their sample-level scores were correlated with the frozen IBDex pathway coordinates.

In macroscopically non-inflamed Crohn’s disease tissue from GSE66207 [26], B1, B2 and B3 separation across the 16-dimensional pathway vector was tested by the pseudo-F statistic

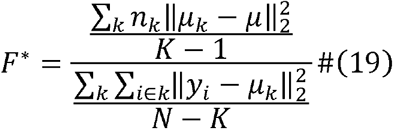

with patient-label permutation. Gene-level programs were also compared with 1,000 size-matched random panels.

GSE171770 supplied 88 serial sigmoid profiles from 16 olamkicept-treated patients [27], including 16 baselines and nine baseline-to-week-14 pairs. Coordinates were expressed relative to each patient’s baseline. Gene-wise follow-up change among responders was compared with the negative frozen inflammation PACP loading across the full panel. Seven responders with complete paired expression contributed at week 14.

Cross-disease transfer used PSC-associated intestinal disease (E-MTAB-9658) and CRC (TCGA-COAD and GSE50760). E-MTAB-9658 contained 9 PSC-associated ulcerative colitis, 11 ulcerative colitis comparator and 12 normal-control patients. The supervised PSC-versus-ulcerative colitis analysis used 9 PSC-associated and 7 comparator patients contributing 21 and 19 biopsies after four comparator patients failed a prespecified quality-control rule. TCGA-COAD used 229 held-out patients. GSE50760 included 18 patients, with a direction fitted in 9 and evaluated in 9. Post-projection phenotype directions were supervised and evaluated by patient-blocked held-out or leave-one-patient-out procedures.

Each cohort used the construction-dataset decoder condition with the most similar mean profile. CRC was queried under right- and left-colon conditions. PSC cecum was mapped to right colon, with rectum and ileum retained. External directions were passed through the unchanged decoder for anatomy-specific PACP. The display UMAP was fitted once to reference latents and only transformed external codes. Cross-disease response reproducibility used the same patient-blocked split-half framework within each anatomical compartment.

### Statistical analysis

Analyses were patient blocked wherever repeated biopsies existed. Empirical permutation P values used (b + 1)/(R + 1), and Benjamini–Hochberg correction [18] was applied within each stated family. Effect sizes accompanied principal P values and bootstrap resampling was patient-level. Split-half distributions were empirical re-estimation references. No sample size was selected after observing an effect. Randomization and blinding were not applicable. Python, NumPy, PyTorch and CUDA seeds were fixed with deterministic resampling offsets.

## Results

### Adversarial conditioning removes cohort identity

IBDex was built from 3,168 biopsies from 1,680 patients across four cohorts, with patients assigned to training, validation or test before model fitting. A training-only audit showed extensive cohort-associated behavior among the 21,912 shared genes. Fixed eligibility gates and reproducibility-weighted ranking produced a 3,000-gene interface without force-retaining any of 213 prespecified landmark genes (Fig. 1a, Table 1, Supplementary Tables 1 and 4–8 and Supplementary Fig. 1). The patient-blocked split contained 2,222 training, 479 validation and 467 test biopsies, so all reported representation metrics were evaluated outside the fitting partition.

**Figure 1.**
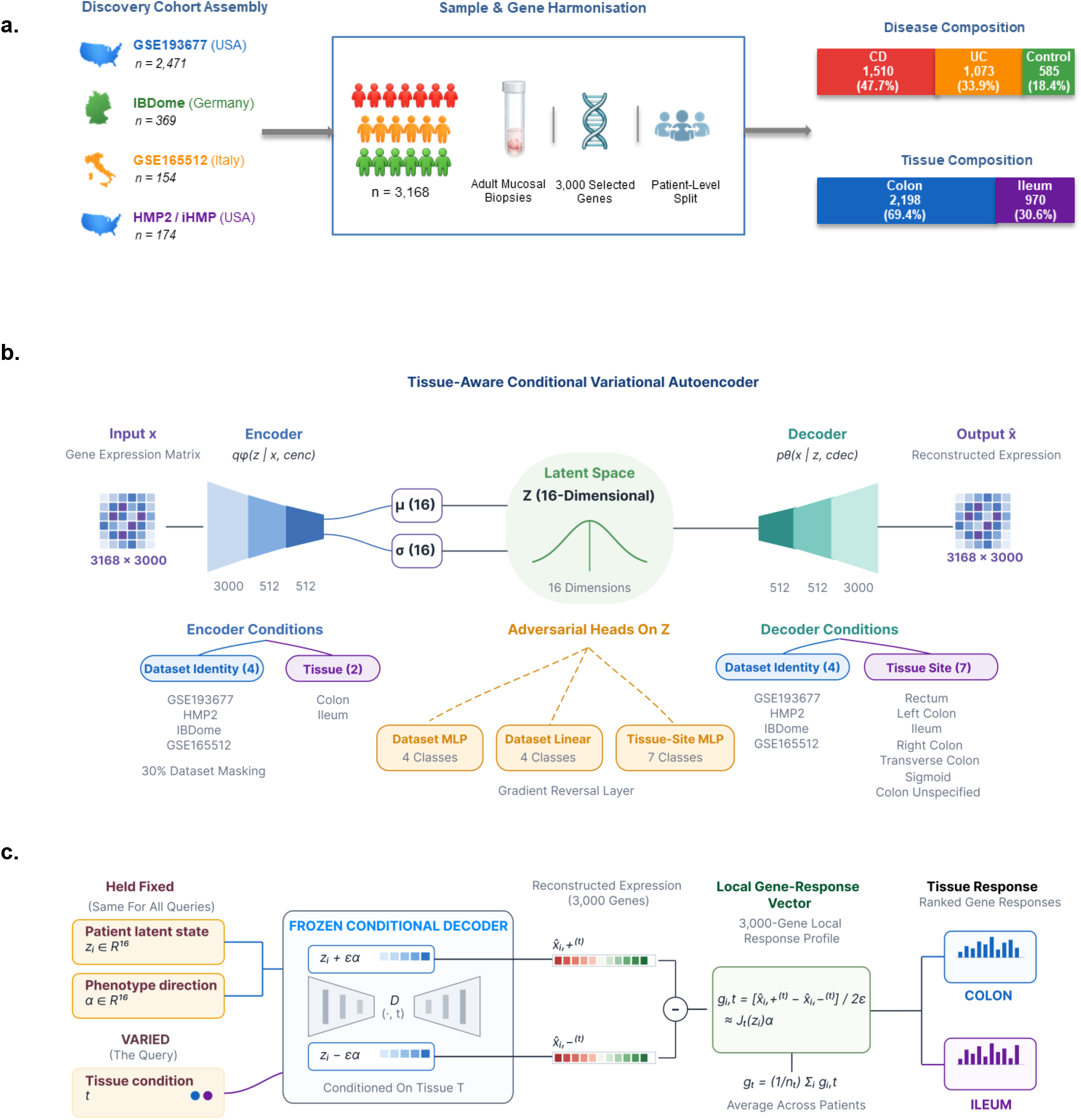
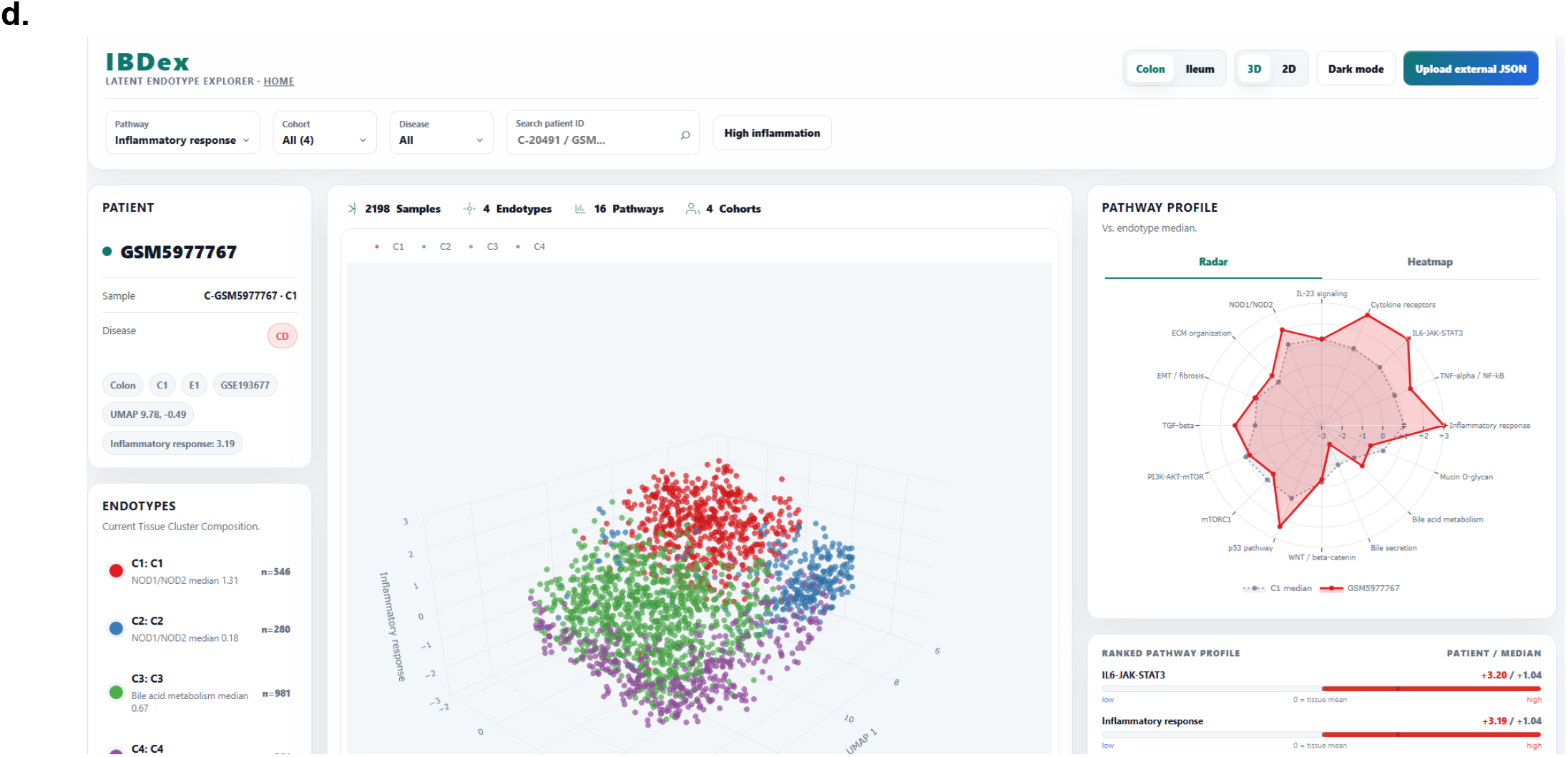
A frozen, tissue-aware reference converts bulk intestinal RNA-seq from cohort-specific analysis into reference-based query. **a**, Four independent adult mucosal biopsy cohorts, GSE193677 (n = 2,471), IBDome (n = 369), GSE165512 (n = 154) and HMP2 (n = 174), were harmonised into a single construction corpus of 3,168 biopsies over a fixed 3,000-gene panel with a patient-level split. The corpus spans Crohn’s disease (n = 1,510, 47.7%), ulcerative colitis (n = 1,073, 33.9%) and non-IBD controls (n = 585, 18.4%) across colon (n = 2,198, 69.4%) and ileum (n = 970, 30.6%). The gene panel, the reference scaling and the partition are fixed once at construction and are never re-estimated for any downstream analysis. **b**, A tissue-aware conditional variational autoencoder maps each expression profile through a 3,000-512-512 encoder into a 16-dimensional latent space conditioned on dataset identity (four classes, 30% dataset masking) and broad tissue. Three adversarial classifier heads attached to the latent code through a gradient-reversal layer, a dataset multilayer perceptron, a dataset linear head and a seven-class tissue-site multilayer perceptron, remove provenance from the coordinates, while a symmetric 512-512-3,000 decoder reconstructs expression conditioned on dataset identity and the full seven-level anatomical site. Deconfounding is therefore applied where the representation is read and anatomy is retained where expression is generated. **c**, The frozen query operator. With the patient latent state z and the phenotype direction alpha both held fixed and only the tissue condition t varied, z + epsilon alpha and z − epsilon alpha are decoded through the same frozen conditional decoder, and the centred difference gives a per-patient local gene-response vector approximating the Jacobian-vector product J(z)alpha, averaged within tissue to the ranked profile used throughout. Because every term is frozen, the response profile for a given phenotype exists before any new cohort is acquired. **d**, Worked single-sample use. One held-out Crohn’s disease colon biopsy (GSM5977767, GSE193677) is placed in the unchanged reference at UMAP coordinates (9.78, −0.49) and returns calibrated pathway scores on the tissue-matched reference scale, including inflammatory response at +3.19 and IL6-JAK-STAT3 at +3.20 relative to the reference median, without any recomputation of the atlas.

**Table 1.**
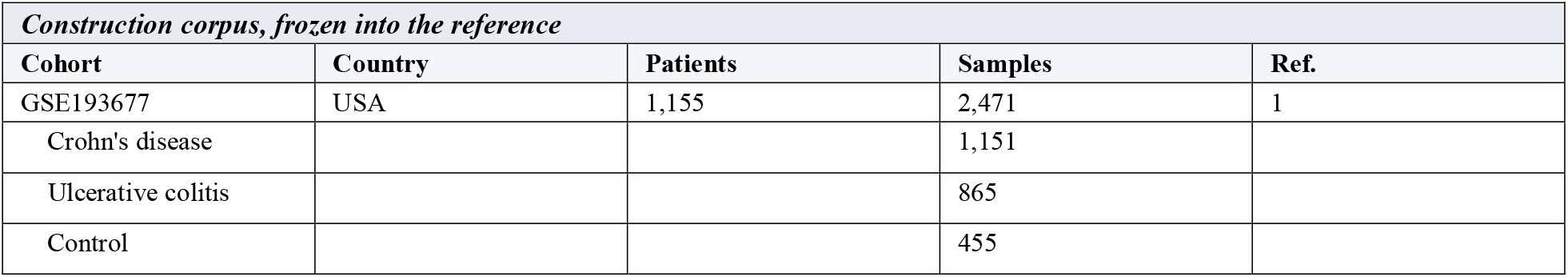
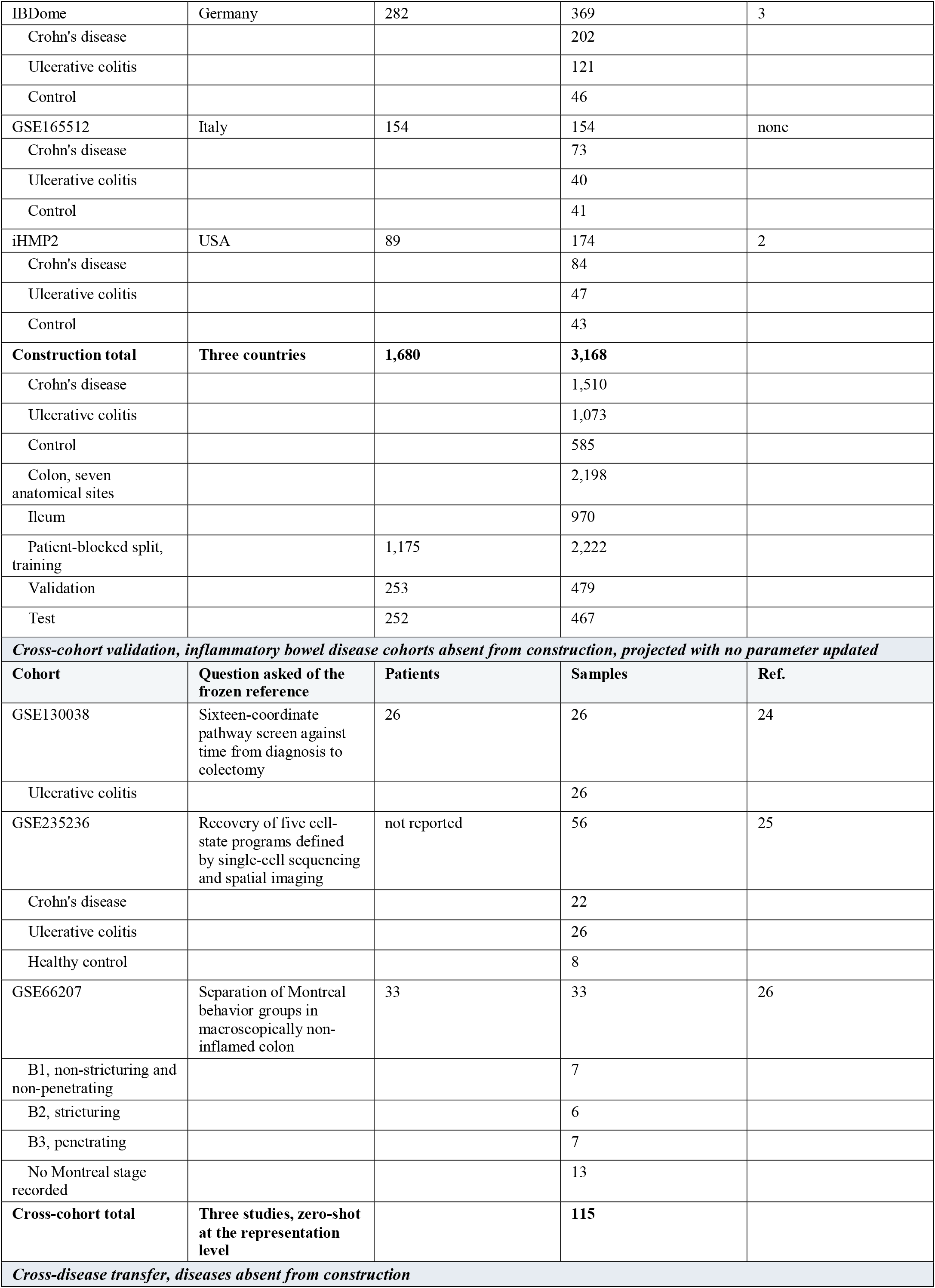
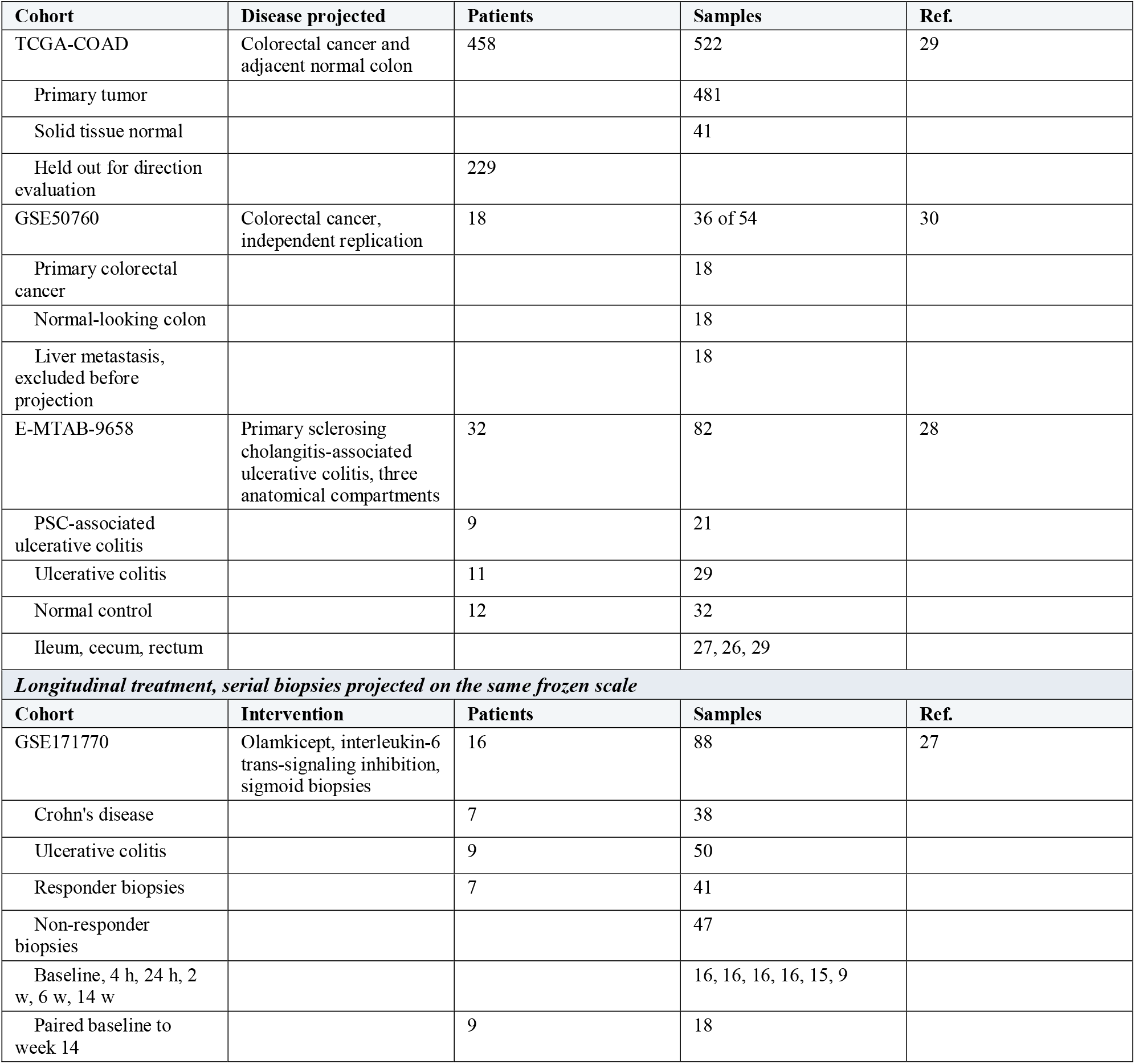
Every cohort the frozen reference was built from, validated against and transferred to, with disease composition given per cohort. Construction counts are biopsies unless stated. No cohort below the construction block contributed to the gene panel, the scaling statistics or any model parameter.

| *Construction corpus, frozen into the reference* |  |  |  |  |
| --- | --- | --- | --- | --- |
| Cohort | Country | Patients | Samples | Ref. |
| GSE193677 | USA | 1,155 | 2,471 | 1 |
| Crohn's disease |  |  | 1,151 |  |
| Ulcerative colitis |  |  | 865 |  |
| Control |  |  | 455 |  |
| IBDome | Germany | 282 | 369 | 3 |
| Crohn's disease |  |  | 202 |  |
| Ulcerative colitis |  |  | 121 |  |
| Control |  |  | 46 |  |
| GSE165512 | Italy | 154 | 154 | none |
| Crohn's disease |  |  | 73 |  |
| Ulcerative colitis |  |  | 40 |  |
| Control |  |  | 41 |  |
| iHMP2 | USA | 89 | 174 | 2 |
| Crohn's disease |  |  | 84 |  |
| Ulcerative colitis |  |  | 47 |  |
| Control |  |  | 43 |  |
| **Construction total** | **Three countries** | **1,680** | **3,168** |  |
| Crohn's disease |  |  | 1,510 |  |
| Ulcerative colitis |  |  | 1,073 |  |
| Control |  |  | 585 |  |
| Colon, seven anatomical sites |  |  | 2,198 |  |
| Ileum |  |  | 970 |  |
| Patient-blocked split, training |  | 1,175 | 2,222 |  |
| Validation |  | 253 | 479 |  |
| Test |  | 252 | 467 |  |
| ***Cross-cohort validation, inflammatory bowel disease cohorts absent from construction, projected with no parameter updated*** |  |  |  |  |
| **Cohort** | **Question asked of the frozen reference** | **Patients** | **Samples** | **Ref.** |
| GSE130038 | Sixteen-coordinate pathway screen against time from diagnosis to colectomy | 26 | 26 | 24 |
| Ulcerative colitis |  |  | 26 |  |
| GSE235236 | Recovery of five cell-state programs defined by single-cell sequencing and spatial imaging | not reported | 56 | 25 |
| Crohn's disease |  |  | 22 |  |
| Ulcerative colitis |  |  | 26 |  |
| Healthy control |  |  | 8 |  |
| GSE66207 | Separation of Montreal behavior groups in macroscopically non-inflamed colon | 33 | 33 | 26 |
| B1, non-stricturing and non-penetrating |  |  | 7 |  |
| B2, stricturing |  |  | 6 |  |
| B3, penetrating |  |  | 7 |  |
| No Montreal stage recorded |  |  | 13 |  |
| **Cross-cohort total** | **Three studies, zero-shot at the representation level** |  | **115** |  |
| ***Cross-disease transfer, diseases absent from construction*** |  |  |  |  |

| Cohort | Disease projected | Patients | Samples | Ref. |
| --- | --- | --- | --- | --- |
| TCGA-COAD | Colorectal cancer and adjacent normal colon | 458 | 522 | 29 |
| Primary tumor |  |  | 481 |  |
| Solid tissue normal |  |  | 41 |  |
| Held out for direction evaluation |  | 229 |  |  |
| GSE50760 | Colorectal cancer, independent replication | 18 | 36 of 54 | 30 |
| Primary colorectal cancer |  |  | 18 |  |
| Normal-looking colon |  |  | 18 |  |
| Liver metastasis, excluded before projection |  |  | 18 |  |
| E-MTAB-9658 | Primary sclerosing cholangitis-associated ulcerative colitis, three anatomical compartments | 32 | 82 | 28 |
| PSC-associated ulcerative colitis |  | 9 | 21 |  |
| Ulcerative colitis |  | 11 | 29 |  |
| Normal control |  | 12 | 32 |  |
| Ileum, cecum, rectum |  |  | 27, 26, 29 |  |
| **Longitudinal treatment, serial biopsies projected on the same frozen scale** |  |  |  |  |
| Cohort | Intervention | Patients | Samples | Ref. |
| GSE171770 | Olamkicept, interleukin-6 trans-signaling inhibition, sigmoid biopsies | 16 | 88 | 27 |
| Crohn's disease |  | 7 | 38 |  |
| Ulcerative colitis |  | 9 | 50 |  |
| Responder biopsies |  | 7 | 41 |  |
| Non-responder biopsies |  |  | 47 |  |
| Baseline, 4 h, 24 h, 2 w, 6 w, 14 w |  |  | 16, 16, 16, 16, 15, 9 |  |
| Paired baseline to week 14 |  | 9 | 18 |  |

PCA and ComBat-seq-style adjustment [16], fitted to the same samples and genes, remained organized by source cohort. By contrast, the tissue-aware CVAE encoded each biopsy into a 16-dimensional latent state while adversarial conditioning suppressed provenance (Figs 1b, 2a and Supplementary Figs 2 and 3).

**Figure 2.**
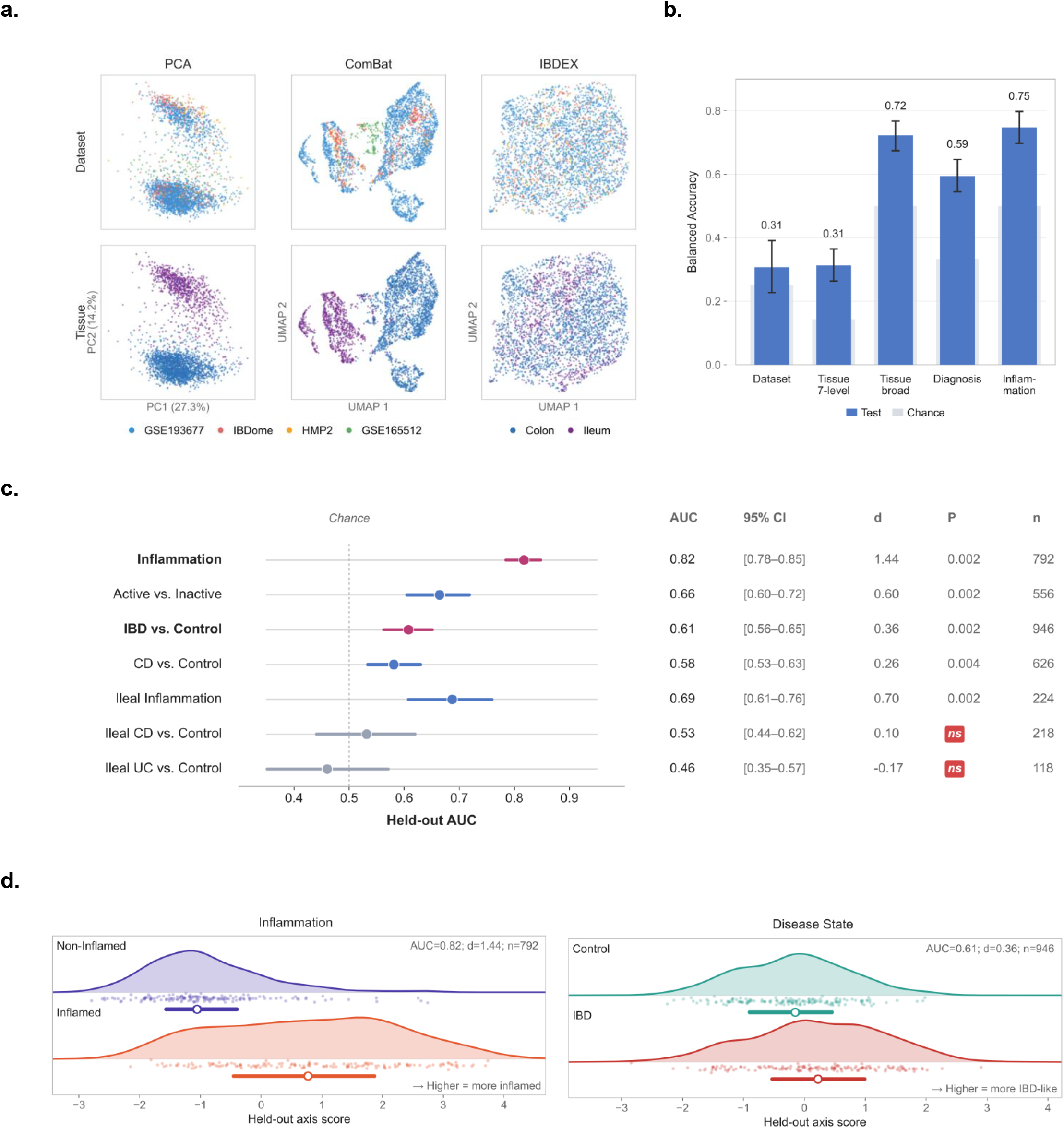
The frozen reference suppresses cohort provenance while retaining anatomical and phenotypic structure. **a**, Principal component analysis of the standardised 3,000-gene panel and UMAP after ComBat adjustment both organise samples by dataset of origin, with cohort structure dominating PC1 and PC2 (27.3% and 14.2% of variance) and persisting after batch correction. The same samples in the frozen IBDEX latent space show no visible dataset organisation. Colouring by broad tissue shows that colon and ileum are geometrically separated in both linear baselines and distributed through the frozen space, where anatomy remains linearly recoverable rather than dominant. **b**, Balanced accuracy of logistic-regression probes decoding each attribute from the frozen latent code on the held-out test partition, each against its own chance level. Dataset identity (0.31) and the seven-level anatomical site (0.31) sit at their chance levels, whereas broad tissue (0.72), diagnosis (0.59) and inflammation (0.75) exceed chance. Provenance is removed to the point of non-decodability while biological structure is preserved. **c**, Held-out discrimination for the seven retained phenotype directions with bootstrap 95% confidence intervals and permutation significance, scored on samples never used to construct the directions. Inflammation reaches AUC 0.82 (95% CI 0.78 to 0.85, d = 1.44, P = 0.002, n = 792) and the inflammation-orthogonal IBD-versus-control direction reaches AUC 0.61 (95% CI 0.56 to 0.65, d = 0.36, P = 0.002, n = 946). The two ileal case-control directions fail permutation validation, are shown in grey and are excluded from interpretation throughout. **d**, Held-out score distributions for the inflammation direction and the orthogonal disease-state direction, split by phenotype group. Both coordinates separate their groups on samples that contributed neither to model training nor to direction construction.

**Figure 3.**
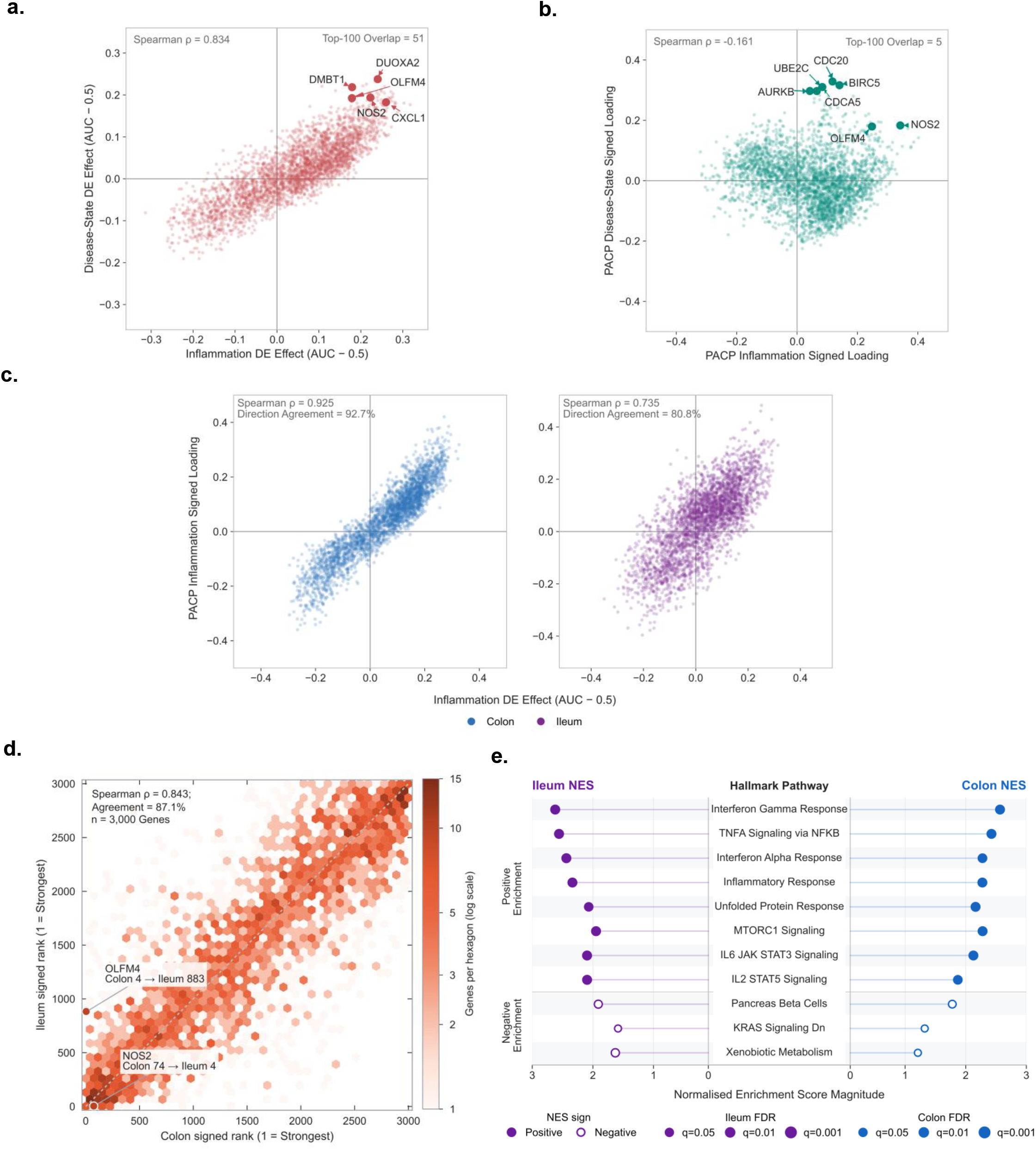
Marginal differential expression conflates inflammation with disease state, whereas the frozen conditional operator separates them. **a**, Marginal differential-expression effect for the inflammation contrast against the marginal effect for the disease-state contrast, both as AUC - 0.5 over the 3,000-gene panel. The two contrasts are close to the same measurement, with signed Spearman rho = 0.834 and 51 of the top 100 genes shared, and both are led by the same epithelial and antimicrobial transcripts *DMBT1, DUOXA2, OLFM4, NOS2* and *CXCL1*. **b**, The same two phenotypes queried as conditional responses through the frozen decoder are close to orthogonal, with signed Spearman rho = −0.161 and only 5 of the top 100 genes shared. The disease-state response resolves a mitotic programme led by *CDC20, UBE2C, AURKB, BIRC5* and *CDCA5* that the marginal contrast does not surface at all. The separation is a property of the estimand rather than of the data, since both estimators were computed on identical samples and identical genes. **c**, Positive control.Where the two estimands do align, the frozen operator recovers the measured contrast, with signed Spearman rho = 0.925 and 92.7% direction agreement in colon and rho = 0.735 with 80.8% agreement in ileum. Inflammation labels were never used to train the representation. **d**, Anatomy reorders a shared programme without reversing it. Colon and ileum signed gene ranks agree at rho = 0.843 with 87.1% direction agreement across all 3,000 genes, while individual genes move substantially, *OLFM4* from rank 4 in colon to 883 in ileum and *NOS2* from rank 74 in colon to 4 in ileum. **e**, Hallmark enrichment of the tissue-specific response rankings recovers interferon gamma response, TNFA signalling via NF-kB, interferon alpha response, inflammatory response, unfolded protein response, MTORC1 signalling, IL6-JAK-STAT3 and IL2-STAT5 signalling as positively enriched in both compartments, with xenobiotic metabolism, KRAS signalling down and pancreas beta cell sets negatively enriched.

Held-out probes showed that provenance suppression preserved biology. Dataset identity reached balanced accuracies of 0.294 in validation and 0.307 in test data against chance 0.250. Median latent-dimension partial R^2^ was 0.0021 and the maximum was 0.0166. In the same representation, colon versus ileum reached 0.743 and 0.723, inflammatory activity 0.777 and 0.747, and diagnosis 0.59 against chance 0.33. A post-training inflammation direction separated inflamed from non-inflamed biopsies at AUC 0.82, 95% confidence interval 0.78–0.85, Cohen’s d = 1.44 and permutation P = 0.002, although inflammation was absent from the training objective (Fig. 2b).

The latent space was continuous rather than divided into stable disease subtypes. Consensus regions were weakly separated and each contained Crohn’s disease, ulcerative colitis and controls from multiple cohorts (Fig. 2c,d, Supplementary Tables 2, 3 and 10–12 and Supplementary Fig. 4). Downstream interpretation therefore used continuous phenotype directions and pathway coordinates. Of 14 candidate phenotype directions, seven survived orthogonalization and five passed held-out permutation testing. Non-validating contrasts were not interpreted (Supplementary Tables 14 and 15). This negative clustering result was consequential, so the atlas was treated as a continuous molecular reference rather than converted into a subtype taxonomy.

**Figure 4.**
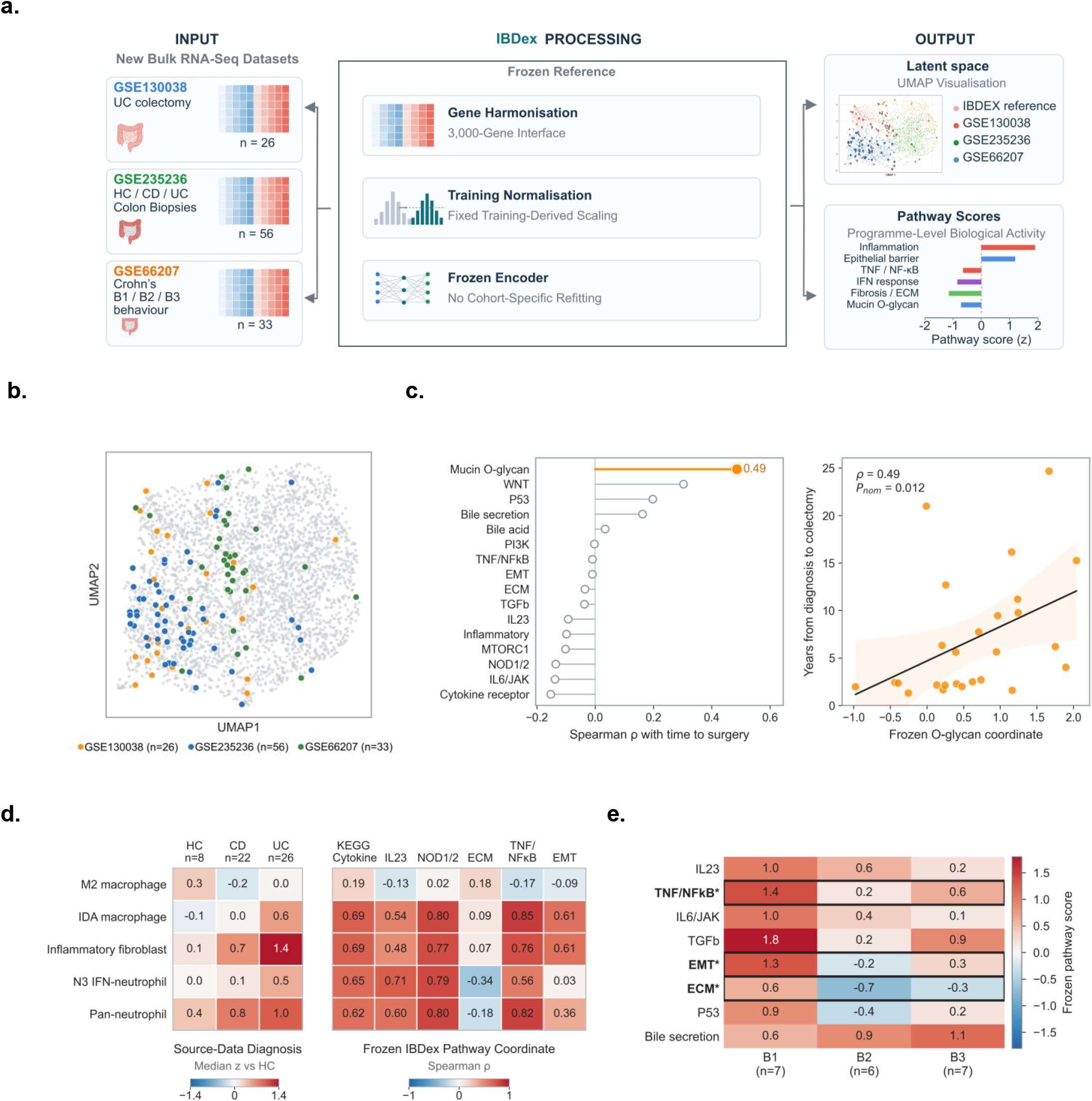
Three IBD cohorts absent from construction are measured against the unchanged reference and recover independently derived biology. **a**, The reference interface. GSE130038 (ulcerative colitis colectomy, n = 26), GSE235236 (healthy control, Crohn’s disease and ulcerative colitis colon biopsies, n = 56) and GSE66207 (Crohn’s disease stratified by Montreal behaviour, n = 33) enter through the fixed 3,000-gene interface and the training-derived scaling and are encoded by the frozen encoder. No cohort-specific refitting, no re-estimation of reference statistics and no retraining are performed at any stage. Cohort sizes are the only measured quantities in this panel; the latent map and the pathway bar chart are schematic and illustrate the output types rather than reporting results. **b**, All 115 projected biopsies fall within the reference manifold, confirming that the frozen encoder places unseen cohorts in the existing coordinate system rather than displacing them to its periphery. **c**, In GSE130038 the frozen mucin O-glycan coordinate is the strongest correlate of time from diagnosis to colectomy among all 16 pathway coordinates (Spearman rho = 0.49, nominal P = 0.012). A coordinate defined before this cohort existed tracks a clinical outcome recorded within it. **d**, In GSE235236 the frozen pathway coordinates align with cell-type signatures derived independently in the source study. The strongest association is between the IDA macrophage signature and the TNF/NF-kB coordinate (rho = 0.85), followed by pan-neutrophil against the same coordinate (0.82), N3 interferon-neutrophil against NOD1/2 (0.79) and inflammatory fibroblast against NOD1/2 (0.77). The M2 macrophage signature reaches no coordinate above 0.19, so the agreement is specific to inflammatory myeloid and fibroblast programmes rather than a general correlation, and the ECM coordinate is negatively associated with both neutrophil signatures. Neither this cohort nor its annotations were exposed to the atlas. **e**, In GSE66207 the frozen coordinates stratify Crohn’s disease behaviour. Groups follow the Montreal classification recorded in the series metadata, giving B1 (n = 7), B2 (n = 6) and B3 (n = 7); one sample carrying a conflicting GEO stage annotation (GSM1616923) is assigned by its Montreal record. The coordinates marked with an asterisk, TNF/NF-kB, EMT and ECM, distinguish the behaviour groups on the fixed pathway scale.

### Frozen pathway coordinates recover tissue biology

Decoder-derived pathway coordinates retained expression-level signal and agreed with independently computed ssGSEA across the 3,168 reference biopsies. Correlations with full-transcriptome ssGSEA had mean ρ = 0.514 and median ρ = 0.550. Panel-matched ssGSEA increased these to 0.559 and 0.604. Among immune pathways, direction agreed in 78.9% of samples and strong opposite-direction calls occurred in 0.03% (Supplementary Table 13 and Supplementary Fig. 5).

**Figure 5.**
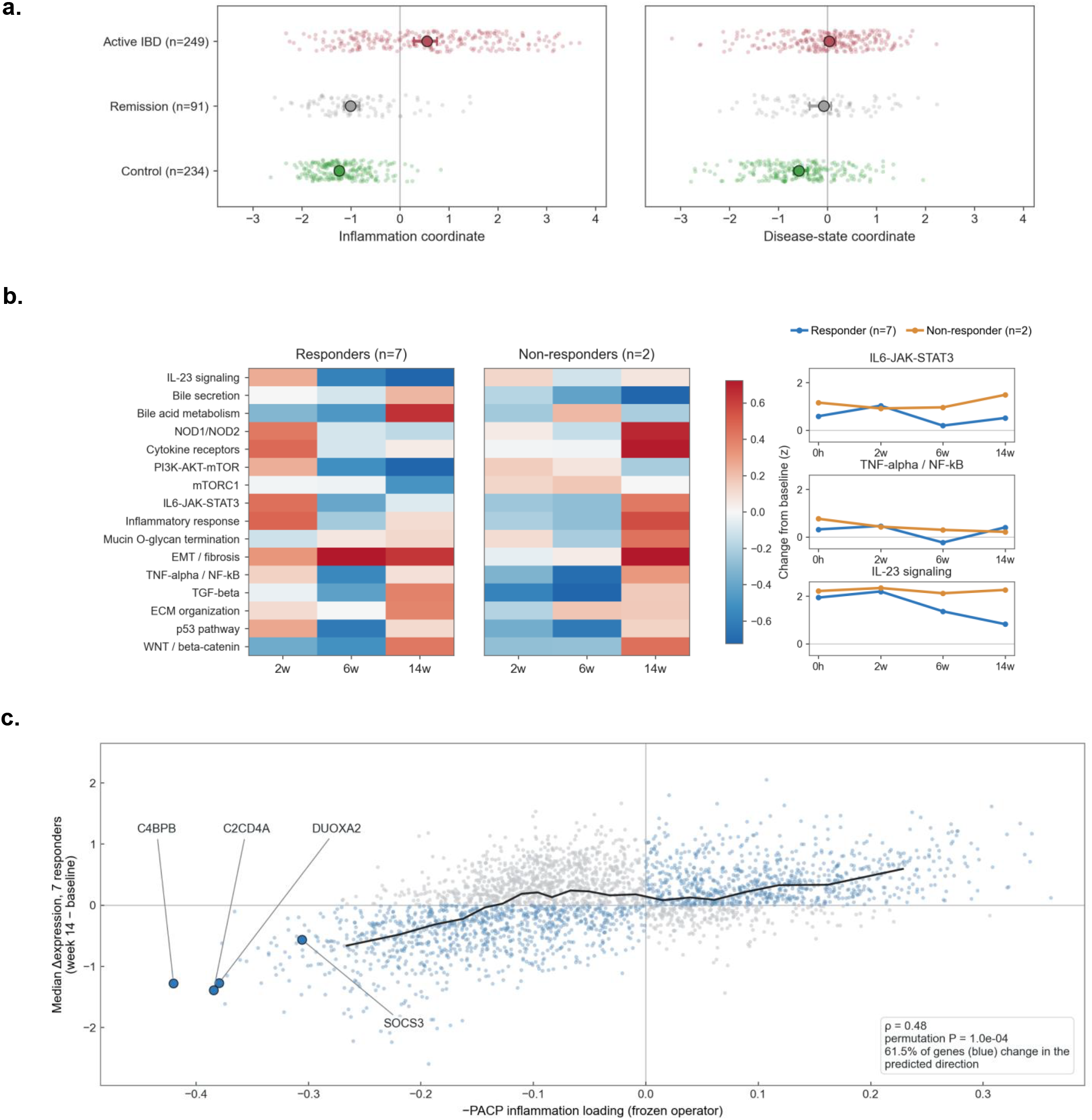
Frozen coordinates separate inflammation resolution from IBD state and anticipate treatment-induced transcriptional change.**a**, Remission resolves the inflammation coordinate but not the Disease-state coordinate. Patient-level frozen coordinates in MSCCR for active IBD (n = 249), concordant remission (n = 91) and control (n = 234); remission approaches control on inflammation (median difference 0.23) but stays at the active-disease level on IBD state (P = 0.16). **b**, Treatment remodels the frozen pathway landscape unevenly over time. Change from baseline in the 16 frozen pathway coordinates at 2, 6 and 14 weeks of olamkicept treatment (GSE171770) in responders (n = 7) and non-responders (n = 9, 8 and 2), with group trajectories for IL6-JAK-STAT3, TNF-alpha / NF-kB and IL-23 signalling. **c**, A response direction learned outside the trial anticipates the genes that change under treatment. Frozen inflammation operator against the median week-14 change in expression across responders for 3,000 genes; ρ = 0.48, permutation P = 1×10^−4^, with 61.5% of genes moving in the predicted direction.

Reliability was coordinate- and compartment-specific. Twelve of fourteen testable coordinates were reliable in colon and eleven in ileum. Bile secretion reached ρ = 0.747 and 0.780, TNF-NF-κB reached 0.733 in colon, and inflammatory response reached 0.767 in ileum. Hallmark bile acid metabolism was unreliable at 0.271 and 0.041. The atlas was not globally collinear, with a median absolute correlation across 105 coordinate pairs of 0.385. Redundancy was concentrated within inflammation, where inflammatory response and IL6-JAK-STAT3 reached |ρ| = 0.99 in colon and were interpreted as related readouts rather than independent discoveries (Supplementary Tables 24 and 25). Two coordinates with insufficient panel coverage were explicitly classified as not testable, preventing sparse gene representation from being mistaken for biological disagreement.

### PACP separates inflammation from disease state

PACP separated biological processes that marginal comparison conflated. It measures the response to movement along a phenotype direction, whereas DE measures the observed difference between groups. Across identical genes and samples, the inflammation and disease-state DE contrasts correlated at ρ = 0.834 and shared 51 of their top 100 genes, including DMBT1, DUOXA2, OLFM4, NOS2 and CXCL1. Their PACP maps instead correlated at ρ = −0.161 and shared five top-100 genes (Fig. 3a,b). Results were insensitive to finite-difference step size and site aggregation, with colon-versus-ileum correlations of 0.839–0.850. The disease-state map exposed mitotic activation and B-cell depletion obscured by the dominant marginal inflammatory contrast (Supplementary Tables 16 and 17 and Supplementary Fig. 6). Because the comparisons used the same biopsies and 3,000 genes, the separation reflected the response estimand rather than a change in samples or feature space.

**Figure 6.**
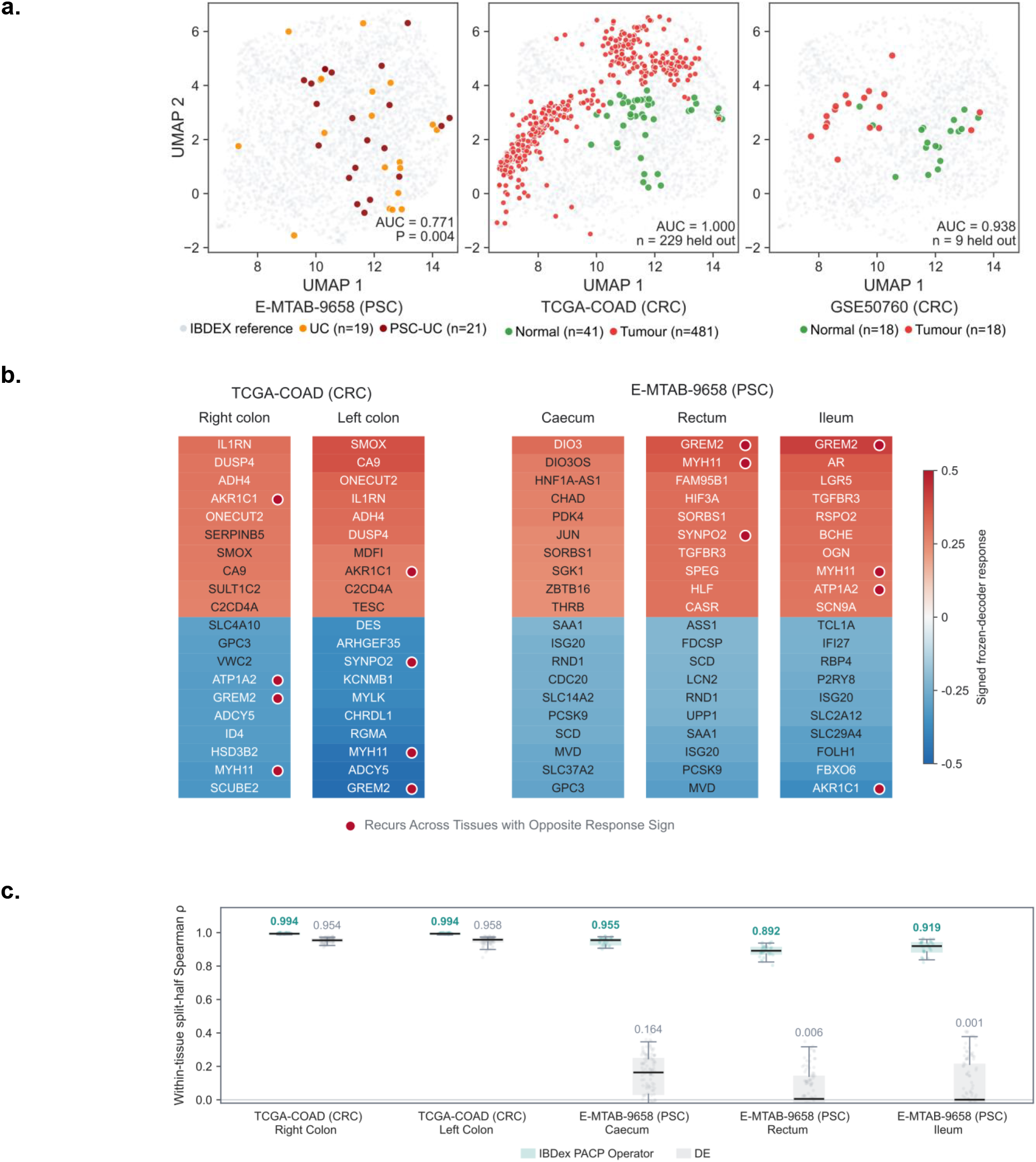
The frozen reference transfers to intestinal diseases absent from construction, and reference-derived rankings remain stable where small-cohort differential expression fails. **a**, Zero-shot projection of three external cohorts into the unchanged reference. PSC-associated colitis (E-MTAB-9658) separates from ulcerative colitis at AUC 0.771 (P = 0.004, n = 19 UC and 21 PSC-UC). Colorectal cancer separates from normal mucosa at AUC 1.000 in TCGA-COAD (n = 229 held out, 41 normal and 481 tumour) and AUC 0.938 in GSE50760 (n = 9 held out, 18 normal and 18 tumour). Projection is zero-shot; the phenotype direction is fitted after projection and never informs the encoder. **b**, Strongest signed frozen-decoder responses within each anatomical compartment. Marked genes recur across compartments with opposite response sign, including *AKR1C1, ATP1A2, GREM2, MYH11* and *SYNPO2*, so the frozen decoder assigns compartment-specific direction rather than replaying one shared programme. **c**, Within-tissue split-half reproducibility of the frozen operator against differential expression estimated de novo, computed on identical samples, contrasts and genes. In the large TCGA-COAD compartments both estimators are reproducible (operator 0.994 against DE 0.954 in right colon, 0.994 against 0.958 in left colon). In the small PSC compartments differential expression collapses (0.164 in caecum, 0.006 in rectum and 0.001 in ileum) while the frozen operator is retained (0.955, 0.892 and 0.919). A reference-derived response therefore remains estimable at sample sizes at which a cohort cannot estimate its own transcriptome-wide ranking, which is the operational case for reference-based analysis of bulk RNA-seq.

Where PACP and DE queried inflammation, they converged at ρ = 0.925 with 92.7% directional agreement in colon and ρ = 0.735 with 80.8% agreement in ileum. Colon and ileum inflammation maps were also broadly aligned at ρ = 0.843 and 87.1% directional agreement, but anatomy sharply re-ranked individual genes. OLFM4 moved from rank 4 in colon to 883 in ileum, whereas NOS2 moved from 74 to 4 (Fig. 3c,d).

Across compartments, 387 of 3,000 genes reversed sign for inflammation and 490 for disease state. No reversal occurred among genes ranked in the top 500 by response magnitude in both compartments for inflammation or the top 1,000 for disease state. Anatomy therefore altered gene usage while preserving the strongest shared responses (Supplementary Table 18).

### Anatomy re-ranks genes but preserves pathway identity

The separation between inflammation and disease state persisted at pathway level. Across 44 Hallmark terms, enrichment profiles were nearly uncorrelated in colon (r = 0.124) and ileum (r = −0.064). TNF-NF-κB reached NES 2.33 and 2.46 on inflammation but −1.59 and −0.87 on disease state. Allograft rejection, IL2-STAT5 and inflammatory response showed similar reversals. Twelve terms reversed sign in colon and fourteen in ileum. Fifteen terms reached FDR < 0.05 on inflammation in each compartment, compared with sixteen in colon and seventeen in ileum on disease state (Fig. 3e and Supplementary Table 26).

Disease state was dominated by G2M checkpoint (NES 3.12 and 3.16) and E2F targets (3.03 and 3.18), both at FDR 0.003. Mitotic spindle reached 2.60 and 2.30, MYC targets 2.15 and 2.20, and type I interferon 2.60 and 2.40. Fatty acid metabolism, glycolysis and cholesterol homeostasis were positive, while xenobiotic metabolism shifted from −1.12 and −1.53 on inflammation to 1.83 and 1.73 on disease state. The residual axis was therefore proliferative, interferon-linked and metabolically reprogrammed, not weaker inflammation.

Despite gene-level re-ranking, colon and ileum pathway profiles correlated at ρ = 0.934 for inflammation and 0.925 for disease state. Every program reaching |NES| ≥ 2 in either compartment retained its sign in the other, nine of nine for inflammation and eight of eight for disease state. Pathway identity was thus more transportable than individual gene rank (Supplementary Table 26).

### Frozen maps remain stable as patient populations change

The frozen response basis remained stable as patient composition changed. Across 30 patient-blocked split halves, PACP rankings were essentially unchanged. Inflammation correlations were approximately 0.999 in colon and ileum, and disease-state correlations were 0.999 and 0.998. DE rankings re-estimated from each patient subset fell to 0.981 and 0.951 for inflammation and 0.954 and 0.849 for disease state. Patients, genes and phenotype definitions were identical, but only PACP reused a fixed response basis. The advantage was greatest in the smaller ileal setting (Supplementary Tables 19 and 20). The fixed atlas therefore supplied a common ranking across resampled cohorts, whereas DE rebuilt the ranking from whichever patients were available.

### Deep remission attenuates inflammation, not disease state

Deep remission attenuated inflammation without normalizing the disease-state axis. Among 93 patients in concordant endoscopic and histological remission, the inflammation coordinate moved to within 0.120 of 241 controls, with AUC 0.58 and P = 0.021. Disease state remained displaced by 0.668, with AUC 0.725 and P = 1.8 × 10^−10^. Relative to 252 patients with concordantly active disease, remission separated strongly on inflammation (AUC 0.165 and P = 1.19 × 10^−21^) but not disease state (AUC 0.518 and P = 0.600) (Fig. 5a) [5]. Thus, the clinical endpoint aligned with inflammatory resolution but did not imply normalization of the broader disease-associated molecular state.

The dissociation persisted across healing definitions. Disease-state displacement from controls was 0.62 for endoscopic remission, 0.63 for histological remission and 0.67 when both agreed. The inflammation gap contracted from 0.40 and 0.59 to 0.12 with concordant remission (Supplementary Table 23). Deep remission therefore substantially attenuated inflammation without restoring a control-like molecular disease state.

### Unseen IBD cohorts recover clinical and cellular structure

The unchanged atlas recovered clinical, cellular and disease-behavior structure across three unseen IBD cohorts comprising 115 biopsies spanning a clinical association in GSE130038, cross-modal cell-state recovery in GSE235236 and disease-behavior structure in macroscopically non-inflamed tissue from GSE66207. All entered the frozen representation without refitting (Fig. 4a,b and Table 1). The three tests asked whether one unchanged coordinate system could recover clinical duration, externally defined cellular programs and disease behavior.

In GSE130038 [24], Reactome mucin O-glycan biosynthesis ranked first among all 16 coordinates against disease duration to colectomy in 26 patients, at ρ = 0.486 and nominal P = 0.012. The same coordinate agreed with full-transcriptome ssGSEA at ρ = 0.507 and P = 0.0082 despite representing only 13 of 26 Reactome genes (Fig. 4c and Supplementary Table 21).

In GSE235236 [25], source-defined cell-state signatures were scored in 56 bulk biopsies. IDA macrophage and pan-neutrophil programs aligned with TNF-NF-κB at ρ = 0.85 and 0.82, while N3 interferon-neutrophil and inflammatory-fibroblast programs aligned with NOD1/2 at 0.79 and 0.77. M2 macrophage reached no coordinate above 0.19. The atlas therefore recovered specific cellular programs from bulk tissue without repeating the source single-cell or spatial analyses (Fig. 4d and Supplementary Table 22).

In macroscopically non-inflamed Crohn’s disease colon from GSE66207 [26], B1 non-stricturing/non-penetrating, B2 stricturing and B3 penetrating disease separated across the frozen pathway space at permutation P = 0.006. TNF-NF-κB, epithelial-mesenchymal transition and extracellular-matrix coordinates contributed (Fig. 4e). Together, the cohorts established clinical, cross-modal and disease-behavior transfer without rebuilding the atlas.

### Treatment response follows pathway-specific kinetics

Frozen coordinates resolved pathway-specific change during treatment. In the independent olamkicept trial [27], seven of nine patients with paired baseline-to-week-14 profiles were responders. In responders, interleukin-23 signaling declined from approximately 2.2 to 0.8, while remaining near 2.2 in two non-responders. PI3K-AKT-mTOR and mTORC1 resolved later, whereas TNF-NF-κB suppression was more transient. These descriptive trajectories show that the fixed atlas resolves pathway-specific kinetics rather than a single before-versus-after response (Fig. 5b). The two non-responders were retained only as descriptive comparators and did not support population-level inference.

The negative frozen inflammation PACP loading also correlated with median week-14-minus-baseline expression change in seven responders at ρ = 0.48 and permutation P = 1.0 × 10^−4^. Concordance emerged by week 6 and peaked at week 14, linking a historical cross-sectional response map to observed change during successful treatment (Fig. 5c).

### Zero-shot projection extends to primary sclerosing cholangitis and colorectal cancer

The representation extended beyond conventional IBD cohorts. PSC-associated intestinal disease and colorectal cancer (CRC) were projected through the unchanged IBD-trained representation. Phenotype directions were fitted only after projection, so representation transfer was zero-shot while disease directions remained supervised (Fig. 6a). In E-MTAB-9658 [28], a post-projection direction separated PSC-associated ulcerative colitis from ulcerative colitis alone under leave-one-patient-out validation at patient-level AUC 0.794. Biopsy-level AUC was 0.627 and fold axes had consensus cosine median 0.982. Across 200 patient-blocked 80/20 splits, biopsy-level AUC ranged from 0.249 to 1.000 around a median of 0.667, reflecting the small cohort of 9 PSC-associated and 7 comparator patients. The wide resampling range emphasizes the uncertainty of biopsy-level estimates in this small external cohort, despite stable patient-level separation and direction concordance.

CRC provided two independent tests. TCGA-COAD [29] reached held-out tumor-normal AUC 1.000 across 229 patients. In GSE50760 [30], an independently derived direction reached AUC 0.938 in 9 held-out patients. Thus, an IBD-trained representation supported separation of a disease absent from construction without rebuilding the latent reference (Fig. 6a).

External directions generated anatomy-specific response maps through the unchanged decoder. CRC response hierarchies differed between right and left colon, and PSC programs differed across cecum, rectum and ileum. AKR1C1, ATP1A2, GREM2, MYH11 and SYNPO2 recurred but changed in magnitude or sign, confirming that anatomical conditioning remained active after disease transfer (Fig. 6b) [28].

Under patient-blocked split halves, CRC response rankings reproduced at ρ = 0.994 in both colon conditions, compared with 0.954 and 0.958 for re-estimated DE. In PSC, DE reproducibility fell to 0.164 in cecum, 0.006 in rectum and 0.001 in ileum, whereas the transferred operator remained at 0.955, 0.892 and 0.919 (Fig. 6c).

The frozen reference therefore supported zero-shot projection, supervised questions within the transferred space and anatomy-conditioned response maps. In each setting, incoming data defined the question but did not alter the representation, pathway scale or decoder used to answer it.

## Discussion

Using IBDex as a fixed patient-scale reference revealed that inflammatory activity and a broader disease-associated molecular state are separable, and that deep remission attenuates the former without normalizing the latter. These findings depended on separating atlas construction from atlas use. Conventional workflows reconstruct normalization, correction, representation and biological contrasts within each incoming cohort. IBDex instead allows historical cohorts to define the gene interface, scaling, encoder, decoder, pathway basis and phenotype-response machinery once, so new samples enter without redefining the molecular scale.

This fixed-reference design remained biologically informative in held-out evaluation. Dataset identity approached chance while anatomy, diagnosis and inflammatory activity remained recoverable. This differs from PCA, which retained dominant cohort covariance, and cohort-level adjustment, which estimates transformations from the datasets being assembled. A reusable atlas must combine provenance suppression with biological interrogability, and the independent probes show that IBDex achieved both.

Within this fixed reference, PACP revealed the response programs underlying the clinical separation. Its distinction from DE is one of estimand. DE measures observed group differences, whereas PACP queries the decoder along a fixed phenotype direction under a specified anatomy. The methods converge for inflammation, where the observed contrast closely matches the queried biology, but diverge when correlated processes are mixed within the groups. PACP separated inflammation from a proliferative, interferon-linked and metabolically altered disease state that marginal DE largely conflated. Because the response operator is reused, its rankings also remained stable when patient composition changed, particularly in the smaller ileal setting. The same fixed coordinate system also transferred beyond the construction cohorts. Three unseen IBD studies recovered a clinical association, cell-state programs originally defined by single-cell and spatial assays, and disease-behavior structure in macroscopically non-inflamed tissue. PSC and colorectal cancer then crossed the disease boundary. Their samples entered the IBD-trained representation without retraining, whereas supervised directions were fitted only after projection and translated by the unchanged decoder into anatomy-specific response maps. New cohorts therefore supplied the biological question while the atlas supplied the coordinate system.

IBDex provided the clearest clinical interpretation by showing that concordant endoscopic and histological remission moved inflammation toward controls but left disease state displaced. Clinical remission and molecular state are therefore separable measurements. The residual axis was enriched for cell-cycle, type I interferon and metabolic programs, whereas inflammation was dominated by TNF and NF-κB signaling. Because both axes used the same frozen genes, decoder and calibration, their separation cannot be attributed to cohort-specific feature selection or normalization. Prospective studies should test whether the residual coordinate predicts relapse, incomplete tissue recovery or later treatment need [5].

Applying the same frozen geometry to longitudinal olamkicept treatment showed that the cross-sectional discoveries remained measurable during intervention. Pathway coordinates followed distinct temporal trajectories, and concordance between the historical inflammation operator and observed responder expression change emerged through treatment [27]. The response direction was not re-estimated in the trial. This supports trial readouts defined before treatment data are analyzed, although the small number of paired responders and non-responders makes these trajectories descriptive rather than confirmatory.

Across these applications, anatomical conditioning remained part of the deployed analytical object rather than merely an optimization aid. The same latent displacement can be queried in colon or ileum, producing compartment-specific gene rankings while preserving pathway identity. This suggests that pathways, rather than individual genes, may be the more transportable biological unit across intestinal compartments. More broadly, conditional generative models can retain clinically meaningful context as an explicit control instead of discarding it as nuisance variation.

Several limitations define the supported interpretation. IBDex complements rather than replaces cell-resolved atlases [8–12] because bulk tissue combines cell abundance with cell-intrinsic regulation. PACP is a directional response of a fitted decoder, not a causal intervention, and external disease directions require labels after projection. The atlas is also restricted to the frozen gene interface, represented anatomical conditions and data compatible with its preprocessing assumptions. Reliability varies by pathway and compartment, so weak or untestable coordinates must remain explicitly flagged rather than interpreted as equivalent measurements.

The decisive next step is prospective validation. Coordinates defined before recruitment should be tested for residual disease, treatment response and relapse across independent trials without within-study re-estimation. Larger longitudinal cohorts can establish which coordinates move reproducibly with intervention and which mark persistent state. Versioned releases should be deliberate and auditable, with declared coordinate reliability and preserved compatibility, so new evidence extends the supported range without silently changing measurements already made.

IBDex establishes a frozen patient-scale atlas that converts historical bulk transcriptomes into a reusable molecular reference. It suppresses cohort provenance while preserving biology, separates response programs obscured by marginal comparison, identifies a disease-state axis that persists after deep remission and supports longitudinal and cross-disease transfer. The broader contribution is not another embedding, but a framework for turning isolated tissue datasets into persistent infrastructure for cross-cohort discovery and prospective validation.

## Data Availability

All construction cohorts are public. GSE193677, GSE165512, GSE130038, GSE235236, GSE66207, GSE50760 and GSE171770 are available from the Gene Expression Omnibus, E-MTAB-9658 from ArrayExpress, iHMP2 from the Inflammatory Bowel Disease Multi'omics Database, IBDome from its project portal and TCGA-COAD from the Genomic Data Commons. Reference coordinates, pathway scores and axis scores for all 3,168 construction biopsies, together with the frozen panel membership and the per-gene standardization and clipping statistics, are distributed with the released package.

https://ibdexmd.org

## Data availability

The frozen IBDex atlas and the complete projection and scoring workflow are publicly available as the Python package ibdex-projector v0.1.0 under an MIT license at github.com/ajithak9945/IBDEX_Package, with an interactive implementation available at ibdexmd.org.

## Acknowledgments

We thank Atharva Waghmare for comments on the manuscript and for assistance with agent development for data extraction and collection.

## Author contributions

A.K.M. conceived the study, designed the reference architecture, assembled and harmonized the cohorts, implemented the model and the phenotype-anchored counterfactual perturbation estimator, performed all analyses, produced the figures and wrote the manuscript. A.A. supervised the project, advised on study design and statistical methodology, and revised the manuscript. All authors read and approved the final manuscript.

## References

1. Argmann C, Hou R, Ungaro RC et al. Biopsy and blood-based molecular biomarker of inflammation in IBD. Gut 2023;72:1271–87. 10.1136/gutjnl-2021-326451

2. Lloyd-Price J, Arze C, Ananthakrishnan AN et al. Multi-omics of the gut microbial ecosystem in inflammatory bowel diseases. Nature 2019;569:655–62. 10.1038/s41586-019-1237-9

3. Plattner C, Sturm G, Kühl AA et al. IBDome: an integrated molecular, histopathological and clinical atlas of inflammatory bowel diseases. Gastroenterology 2026. 10.1053/j.gastro.2026.05.023

4. Kennedy NA, Heap GA, Green HD et al. Predictors of anti-TNF treatment failure in anti-TNF-naive patients with active luminal Crohn’s disease: a prospective, multicentre, cohort study. Lancet Gastroenterol Hepatol 2019;4:341–53. 10.1016/S2468-1253(19)30012-3

5. Turner D, Ricciuto A, Lewis A et al. STRIDE-II: an update on the Selecting Therapeutic Targets in Inflammatory Bowel Disease (STRIDE) initiative of the International Organization for the Study of IBD. Gastroenterology 2021;160:1570–83. 10.1053/j.gastro.2020.12.031

6. West NR, Hegazy AN, Owens BMJ et al. Oncostatin M drives intestinal inflammation and predicts response to tumor necrosis factor-neutralizing therapy in patients with inflammatory bowel disease. Nat Med 2017;23:579–89. 10.1038/nm.4307

7. Martin JC, Chang C, Boschetti G et al. Single-cell analysis of Crohn’s disease lesions identifies a pathogenic cellular module associated with resistance to anti-TNF therapy. Cell 2019;178:1493–508.e20. 10.1016/j.cell.2019.08.008

8. Regev A, Teichmann SA, Lander ES et al. The Human Cell Atlas. eLife 2017;6:e27041. 10.7554/eLife.27041

9. Lopez R, Regier J, Cole MB et al. Deep generative modeling for single-cell transcriptomics. Nat Methods 2018;15:1053–8. 10.1038/s41592-018-0229-2

10. Lotfollahi M, Naghipourfar M, Luecken MD et al. Mapping single-cell data to reference atlases by transfer learning. Nat Biotechnol 2022;40:121–30. 10.1038/s41587-021-01001-7

11. Lotfollahi M, Wolf FA, Theis FJ. scGen predicts single-cell perturbation responses. Nat Methods 2019;16:715–21. 10.1038/s41592-019-0494-8

12. Nie H, Lin P, Zhang Y et al. Single-cell meta-analysis of inflammatory bowel disease with scIBD. Nat Comput Sci 2023;3:522–31. 10.1038/s43588-023-00464-9

13. Sohn K, Yan X, Lee H. Learning structured output representation using deep conditional generative models. Adv Neural Inf Process Syst 2015;28:3483–91.

14. Milacic M, Beavers D, Conley P et al. The Reactome Pathway Knowledgebase 2024. Nucleic Acids Res 2024;52:D672–8. 10.1093/nar/gkad1025

15. Kanehisa M, Furumichi M, Sato Y et al. KEGG for taxonomy-based analysis of pathways and genomes. Nucleic Acids Res 2023;51:D587–92. 10.1093/nar/gkac963

16. Zhang Y, Parmigiani G, Johnson WE. ComBat-seq: batch effect adjustment for RNA-seq count data. NAR Genom Bioinform 2020;2:lqaa078. 10.1093/nargab/lqaa078

17. Ganin Y, Ustinova E, Ajakan H et al. Domain-adversarial training of neural networks. J Mach Learn Res 2016;17:1–35.

18. Benjamini Y, Hochberg Y. Controlling the false discovery rate: a practical and powerful approach to multiple testing. J R Stat Soc Ser B 1995;57:289–300. 10.1111/j.2517-6161.1995.tb02031.x

19. Monti S, Tamayo P, Mesirov J et al. Consensus clustering: a resampling-based method for class discovery and visualization of gene expression microarray data. Mach Learn 2003;52:91–118. 10.1023/A:1023949509487

20. Traag VA, Waltman L, van Eck NJ. From Louvain to Leiden: guaranteeing well-connected communities. Sci Rep 2019;9:5233. 10.1038/s41598-019-41695-z

21. Liberzon A, Birger C, Thorvaldsdóttir H et al. The Molecular Signatures Database hallmark gene set collection. Cell Syst 2015;1:417–25. 10.1016/j.cels.2015.12.004

22. Subramanian A, Tamayo P, Mootha VK et al. Gene set enrichment analysis: a knowledge-based approach for interpreting genome-wide expression profiles. Proc Natl Acad Sci USA 2005;102:15545–50. 10.1073/pnas.0506580102

23. Barbie DA, Tamayo P, Boehm JS et al. Systematic RNA interference reveals that oncogenic KRAS-driven cancers require TBK1. Nature 2009;462:108–12. 10.1038/nature08460

24. Eshelman MA, Jeganathan NA, Schieffer KM et al. Elevated colonic mucin expression correlates with extended time to surgery for ulcerative colitis patients. J Gastrointestin Liver Dis 2019;28:405–13. 10.15403/jgld-250

25. Garrido-Trigo A, Corraliza AM, Veny M et al. Macrophage and neutrophil heterogeneity at single-cell spatial resolution in human inflammatory bowel disease. Nat Commun 2023;14:4506. 10.1038/s41467-023-40156-6

26. Peck BCE, Weiser M, Lee SE et al. MicroRNAs classify different disease behavior phenotypes of Crohn’s disease and may have prognostic utility. Inflamm Bowel Dis 2015;21:2178–87. 10.1097/MIB.0000000000000478

27. Schreiber S, Aden K, Bernardes JP et al. Therapeutic interleukin-6 trans-signaling inhibition by olamkicept in patients with active inflammatory bowel disease. Gastroenterology 2021;160:2354–66.e11. 10.1053/j.gastro.2021.02.062

28. Ilott NE, Neyazi M, Arancibia-Cárcamo CV et al. Tissue-dependent transcriptional and bacterial associations in primary sclerosing cholangitis-associated inflammatory bowel disease. Wellcome Open Res 2021;6:199. 10.12688/wellcomeopenres.16901.2

29. Cancer Genome Atlas Network. Comprehensive molecular characterization of human colon and rectal cancer. Nature 2012;487:330–7. 10.1038/nature11252

30. Kim S-K, Kim S-Y, Kim J-H et al. A nineteen gene-based risk score classifier predicts prognosis of colorectal cancer patients. Mol Oncol 2014;8:1653–66. 10.1016/j.molonc.2014.06.016

