## supplementary for "IBDex: A Frozen Patient-Scale Atlas for Bulk Transcriptomics in Inflammatory Bowel Disease"

### Abbreviations

Supplementary Table A | Abbreviations used throughout this document.

| **Abbreviation** | **Definition** |
|---|---|
| AUC | area under the receiver operating characteristic curve |
| CD | Crohn's disease |
| ComBat-seq | per-dataset batch adjustment for sequencing count data |
| CRC | colorectal cancer |
| CVAE | conditional variational autoencoder |
| DE | differential expression |
| ECM | extracellular matrix |
| EMT | epithelial-to-mesenchymal transition |
| FDR | false discovery rate |
| GRL | gradient-reversal layer |
| GSEA | gene set enrichment analysis |
| IBD | inflammatory bowel disease |
| IBDex | IBD Explorer the frozen intestinal reference described in this work |
| LOPO | leave-one-patient-out |
| NES | normalized enrichment score |
| PACP | phenotype-anchored counterfactual perturbation |
| PCA | principal component analysis |
| PSC | primary sclerosing cholangitis |
| ssGSEA | single-sample gene set enrichment analysis |
| UC | ulcerative colitis |
| UMAP | uniform manifold approximation and projection |
| rho | Spearman rank correlation coefficient |

### Supplementary Tables

Supplementary Table 1 | Construction corpus and patient-blocked partition. No patient contributes samples to more than one partition, and gene selection, reference scaling and model fitting used the training partition alone. The partition is the reason every number in this document is a held-out number rather than a fit statistic.

| Partition | Patients | Samples | Role |
|---|---|---|---|
| Training | 1,175 | 2,222 | gene panel, scaling, model fitting |
| Validation | 253 | 479 | checkpoint selection only |
| Test | 252 | 467 | representation probes and phenotype validation |
| Total | 1,680 | 3,168 | |

Supplementary Table 2 | Held-out information probes on the frozen latent codes. Balanced accuracy against each target's own chance level. Probes were fitted after freezing and played no part in model selection. Inflammatory activity reaches 0.777 and 0.747 without ever entering the training objective, while dataset identity stays within 0.057 of chance, which is the regime a reusable reference requires.

| Target | Classes | Chance | Validation | Test |
|---|---|---|---|---|
| Dataset identity | 4 | 0.250 | 0.294 | 0.307 |
| Anatomical site, seven-level | 7 | 0.143 | 0.318 | 0.313 |
| Diagnosis | 3 | 0.333 | 0.507 | 0.594 |
| Inflammatory activity | 2 | 0.500 | 0.777 | 0.747 |

Supplementary Table 3 | Per-dimension variance partitioning of the frozen latent space. Dataset provenance is distributed rather than concentrated, with a median partial R-squared of 0.0021 across all 16 dimensions. Full per-dimension values are in the source data.

| Quantity | Median | Maximum | Dimension at maximum |
|---|---|---|---|
| Partial R-squared, dataset | 0.0021 | 0.0166 | 12 |
| Partial R-squared, anatomical site | 0.0201 | 0.1093 | 12 |
| Partial R-squared, diagnosis | 0.0121 | 0.0719 | 1 |
| Full R-squared | 0.0319 | 0.1689 | 12 |

Supplementary Table 4 | Cohort-compatibility audit of the shared transcriptome, training partition only. Roughly one gene in five measured in every cohort carries study identity rather than intestinal biology, which is the quantitative case for fixing the interface before fitting the model.

| Class | Genes | Share | Interpretation |
|---|---|---|---|
| Cohort-compatible | 12,777 | 58.3% | behavior consistent across the four cohorts |
| Retained for review | 4,695 | 21.4% | partly cohort-associated behavior |
| Cohort-barcode-like | 4,165 | 19.0% | study of origin predicts value better than biology |
| Presence-absence artifact | 275 | 1.3% | detected in some cohorts, absent in others |
| Shared transcriptome | 21,912 | 100% | intersection across all four cohorts |

Supplementary Table 5 | Eligibility gates applied before ranking. Gates are hard, so no gene could score its way past a technical failure, with one declared exception: 122 genes falling just below the expression floor were readmitted only on meeting a stricter combined standard for detection, cross-cohort reproducibility and both cohort-confounding measures.

| Gate | Requirement | Rationale |
|---|---|---|
| Expression support | sufficient expression level | avoid quantification noise |
| Detection frequency | sufficient detection across samples | avoid sparse features |
| Global dataset eta squared | below threshold | exclude cohort-driven genes |
| Within-tissue dataset eta squared | below threshold | exclude cohort effects masked by anatomy |
| Presence-absence behavior | absent | exclude platform artifacts |
| Sex-linked genes | excluded | encode donor characteristics, not mucosal state |
| Mitochondrial genes | excluded | reflect dissociation and perfusion |
| Hemoglobin genes | excluded | reflect blood content of the biopsy |
| Immunoglobulin variable regions | constrained | prevent clonotype-dominated features occupying the panel |
| Genes passing all gates | 6,058 | eligible for ranking |

Supplementary Table 6 | Composite ranking score and its maximum contributions, fixed before scoring. Reproducibility outweighs disease association, which is the opposite of a biomarker-discovery weighting and is deliberate for a reference panel.

| Component | Measures | Maximum points |
|---|---|---|
| S_endo | residual biological heterogeneity after adjusting for diagnosis, tissue and dataset | 40 |
| S_repro | cross-cohort concordance of that heterogeneity | 20 |
| S_dis | disease association | 15 |
| S_safety | penalty for batch-driven and tissue-driven behavior | 10 |
| S_ext | independent external biological evidence | 15 |
| Total | additive across components | 100 |

Supplementary Table 7 | External evidence term and its fixed source weights. The term is capped so that annotation cannot dominate measured behavior, because annotation databases are themselves biased toward genes already studied in this disease.

| Source | Weight | Evidence type |
|---|---|---|
| STRING | 0.20 | protein-protein association network centrality |
| Reactome | 0.20 | curated pathway membership |
| KEGG | 0.15 | curated pathway membership |
| Open Targets | 0.15 | target-disease association and tractability |
| Gene Ontology Biological Process | 0.10 | functional annotation |
| NHGRI-EBI GWAS Catalog | 0.10 | genetic risk association |
| Prespecified landmark set | 0.10 | mechanism and therapeutic relevance |

Supplementary Table 8 | Landmark audit. All 213 mechanism and therapeutic landmark genes were screened against the ranked panel and none was force-added. The 53 that did not reach the top 3,000 were held out by gates fixed before ranking, and each is recorded with the gate it failed.

| Item | Value |
|---|---|
| Landmark genes screened against the ranked panel | 213 |
| Entered the panel on composite score alone | 160 |
| Held out by the eligibility gates | 53 |
| Of those, failing a batch or tissue-confounding gate | 47 |
| Of those, failing on expression or detection alone | 6 |
| Of those, otherwise eligible and still held out | 33 |
| Genes force-added to the panel after the gates | 0 |
| Final panel size | 3,000 |
| Independent check | None of the 53 appears as a column of the training matrix or in the normalization statistics shipped with the released package |

Supplementary Table 9 | Selected model configuration, frozen at the end of construction. The search optimized validation total loss only. Search ranges are held in the study record and are inserted where marked. Fixing one completed search rather than selecting among repeated searches is what prevents a downstream biological result from choosing the architecture.

| Parameter | Selected | Search range |
|---|---|---|
| Latent dimension | 16 | 16 or 32 |
| Hidden width | 512 | 128, 256 or 512 |
| Depth | 2 | 1, 2 or 3 |
| Dropout | 0.10 | 0.1 to 0.4 |
| Batch size | 32 | 32, 64 or 128 |
| Learning rate | $6 \times 10^{-4}$ | $1 \times 10^{-4}$ to $1 \times 10^{-3}$ |
| Weight decay | $3.1 \times 10^{-4}$ | not searched, set at $3.1 \times 10^{-4}$ |
| beta maximum | 1 | 0.5 to 2.0 |
| beta cycle length | 1 | fixed |
| beta plateau fraction | 1.0 | fixed |
| Reconstruction weight | 1 | fixed |
| Dataset sampler weight | 1.0 | fixed |
| Tissue sampler weight | 0.25 | fixed |
| Balanced dataset batches | true | fixed |
| lambda, dataset adversary | 120 | 120 or 140 |
| lambda, linear dataset adversary | 120 | 120 or 140 |
| lambda, tissue-site adversary | 80 | 60, 80 or 100 |
| beta ramp, epochs | 1 to 120 | fixed |
| Adversary ramp, epochs | 1 to 120 | fixed |

| | | |
|---|---|---|
| Tissue adversary ramp, epochs | 1 to 120 | fixed |
| Encoder condition dimension | 6 | fixed by design |
| Decoder condition dimension | 11 | fixed by design |
| Selected checkpoint | epoch 179 | minimum validation total |
| Validation total at checkpoint | 1021.4342 | selection criterion |
| Early stopping | epoch 209 | patience 30, no checkpoint before epoch 150 |

Supplementary Table 10 | Base partitions generated for consensus clustering. Breadth is deliberate, so that a negative result cannot be attributed to a single algorithm failing to find structure another would recover.

| **Family** | **Parameter grid** | **Repeats** |
|---|---|---|
| k-means | k = 2 to 8 | 50 initializations |
| Gaussian mixture | k = 2 to 8, full, diagonal and tied covariance | 10 initializations |
| Spectral | neighborhood sizes 10, 15, 25 | k = 2 to 8 |
| Leiden | neighborhood sizes 10, 15, 25, multiple resolutions | latent k-nearest-neighbor graph |
| Scopes | colon, ileum, tissue-mixed reference | minimum 30 samples |
| Selection indices | silhouette, Calinski-Harabasz, Davies-Bouldin | |
| Size constraints | each region at least 3% of scope and 8 samples, none above 70% | |

Supplementary Table 11 | Consensus regions recovered within each compartment and their dominant programs. Reported as reproducible topology of the frozen space, not as a proposed subtype taxonomy. Every region contains all three diagnoses and draws from more than one construction cohort.

| **Region** | **Compartment** | **n** | **Dominant program** | **Supporting markers** |
|---|---|---|---|---|
| C1 | Colon | 546 | Inflammatory and remodeling | DMBT1, OLFM4, PSMB9 |
| C2 | Colon | 280 | Low-inflammatory, bile-acid-leaning | - |
| C3 | Colon | 981 | WNT and epithelial, low inflammation | - |
| C4 | Colon | 391 | Bile-acid epithelial with stress and proliferation | - |
| I1 | Ileum | 227 | Inflammatory, immune dominant | LCN2, DUOX2, NOS2 |
| I2 | Ileum | 427 | Bile-acid metabolic, low inflammation | - |
| I3 | Ileum | 56 | Fibrotic and remodeling, WNT and TGF character | - |
| I4 | Ileum | 260 | Inflammatory with stress and remodeling | - |

Supplementary Table 12 | Consensus stability against a 250-permutation co-membership null. Values in this range are what a continuum with local density variation produces. Discrete subtypes would approach one and unstructured noise would approach zero.

| **Statistic** | **Colon** | **Ileum** | **Reading** |
|---|---|---|---|
| Adjusted Rand index | 0.416 | 0.460 | moderate agreement across the base partitions |
| Jaccard index | 0.401 | 0.439 | moderate region membership stability |
| Consensus gap against null | 0.401 | 0.437 | structure exceeds the permutation null |
| Between-region silhouette | weak | weak | regions are not separable |
| Regions containing CD, UC and control | all | all | no region is a diagnosis |
| Regions drawing from more than one cohort | all | all | no region is a cohort |

Supplementary Table 13 | Agreement between frozen pathway coordinates and expression-derived ssGSEA across all 3,168 reference biopsies. Values are Spearman correlations summarised across the fourteen coordinates testable in both compartments, the remaining two having gene sets too sparsely covered to score. The panel-matched comparison isolates the cost of the fixed gene interface. Direction agreement and opposite-direction calls are computed on scores standardised within compartment across the five testable immune coordinates, and a call counts as strong when both standardised scores exceed 1.5 in absolute value with opposite signs.

| Comparison | Mean rho | Median rho |
|---|---|---|
| ssGSEA over the full measured transcriptome | 0.496 | 0.521 |
| ssGSEA restricted to the 3,000-gene interface | 0.584 | 0.631 |
| Immune-set direction agreement, panel-matched | 76.1% | |
| Strong opposite-direction calls | 0.03% | |

Supplementary Table 14 | All 14 candidate phenotype directions with group sizes and unprojected norms. Candidates were generated only where both groups contained at least 25 training samples.

| Candidate direction | n group A | n group B | Norm |
|---|---|---|---|
| Inflammation | 613 | 1,240 | 1.615 |
| Active vs inactive | 196 | 1,067 | 0.960 |
| IBD vs control | 1,809 | 413 | 1.258 |
| CD vs control | 1,056 | 413 | 1.306 |
| UC vs control | 753 | 413 | 1.300 |
| CD vs UC | 1,056 | 753 | 0.692 |
| Ileal inflammation | 167 | 359 | 1.579 |
| Ileal CD vs control | 391 | 128 | 1.619 |
| Ileal UC vs control | 162 | 128 | 0.653 |
| Ileal CD vs UC | 391 | 162 | 1.162 |
| Colonic inflammation | 446 | 881 | 1.744 |
| Colonic CD vs control | 665 | 285 | 1.195 |
| Colonic UC vs control | 591 | 285 | 1.540 |
| Colonic CD vs UC | 665 | 591 | 0.917 |

Supplementary Table 15 | Orthogonalization residuals and held-out validation. Fourteen candidates reduce to seven independent directions and then to five that survive permutation testing. Residual norm is relative to the unprojected contrast.

| Direction | Residual norm | Retained | Held-out AUC | Permutation P |
|---|---|---|---|---|
| Inflammation | 1.000 | yes | 0.818 | 0.002 |
| Active vs inactive | 0.658 | yes | 0.664 | 0.002 |
| IBD vs control | 0.460 | yes | 0.608 | 0.002 |
| CD vs control | 0.216 | yes | 0.581 | 0.004 |
| Ileal inflammation | 0.482 | yes | 0.687 | 0.002 |
| Ileal CD vs control | 0.246 | yes | 0.532 | 0.511, not significant |
| Ileal UC vs control | 0.402 | yes | 0.460 | 0.403, not significant |
| UC vs control | $4.1 \times 10^{-7}$ | no | | |
| CD vs UC | $8.6 \times 10^{-7}$ | no | | |
| Ileal CD vs UC | $5.9 \times 10^{-8}$ | no | | |
| Colonic inflammation | 0.0061 | no | | |
| Colonic CD vs control | 0.0298 | no | | |
| Colonic UC vs control | 0.0292 | no | | |
| Colonic CD vs UC | 0.0878 | no | | |

Supplementary Table 16 | Step-size and aggregation sensitivity of the response operator, on signed ranks. The reported map is unchanged by two orders of magnitude of step size and survives both alternative site-aggregation rules, which is what licenses treating the colon-to-ileum separation as decoder geometry rather than as a numerical or sampling artifact. The two reference rows differ slightly because the aggregation check recomputes both compartment operators from the stored site Jacobians.

| Setting | Reported choice | Alternative tested | Signed rank correlation with the reported map | Signed colon-versus-ileum correlation |
|---|---|---|---|---|
| Finite-difference step | 1e-2 | reference | 1.0000 | 0.8425 |
| Finite-difference step | 1e-2 | 1e-3 | 1.0000 | 0.8425 |
| Finite-difference step | 1e-2 | 1e-1 | 1.0000 | 0.8426 |
| Site aggregation | sample-count weighted | reference | 1.0000 | 0.8453 |
| Site aggregation | sample-count weighted | equal site weights | 0.9956 | 0.8500 |
| Site aggregation | sample-count weighted | elementwise median | 0.9941 | 0.8393 |

Supplementary Table 17 | Site-level Jacobian estimation and compartment aggregation. Compartment operators are sample-count weighted combinations of their constituent sites. Reporting site-level operators before aggregation is what makes the compartment operator auditable against its constituent sites.

| Anatomical site | Samples | Compartment |
|---|---|---|
| Rectum | 961 | colon |
| Right colon | 498 | colon |
| Left colon | 292 | colon |
| Sigmoid colon | 203 | colon |
| Transverse colon | 113 | colon |
| Colon unspecified | 131 | colon |
| Ileum | 970 | ileum |

Supplementary Table 18 | Sign reversal between compartments is confined to low-magnitude genes. Reversals concentrate in the weakly responding tail, and no gene reverses while ranking in the top 500 by magnitude in both compartments. Reversal rate is stratified by the better of the two magnitude ranks, for the inflammation direction.

| Quantity | Inflammation | IBD vs control |
|---|---|---|
| Signed rank correlation, colon vs ileum | 0.843 | 0.858 |
| Magnitude rank correlation | 0.575 | 0.644 |
| Genes keeping direction | 2,613 | 2,510 |
| Genes reversing direction | 387 | 490 |
| Direction agreement | 87.1% | 83.7% |
| Reversals with both magnitude ranks in the top 500 | 0 | 0 |
| Reversals with both magnitude ranks in the top 1,000 | 1 | 0 |
| Median of the smaller loading, reversed genes | 0.022 | 0.011 |
| Median of the smaller loading, same-direction genes | 0.090 | |
| Signed correlation after removing all reversed genes | 0.884 | |
| Reversal rate, better magnitude rank 1 to 100 | 0.6% | |
| Reversal rate, better magnitude rank 101 to 300 | 2.4% | |
| Reversal rate, better magnitude rank 301 to 1,000 | 5.1% | |
| Reversal rate, better magnitude rank 1,001 to 2,000 | 12.6% | |
| Reversal rate, better magnitude rank 2,001 to 3,000 | 33.1% | |

Supplementary Table 19 | Patient-blocked split-half reproducibility within the construction corpus, over 30 draws. Signed rank correlation between the two half rankings. Intervals are percentile intervals over the 30 draws, the 2.5th to 97.5th for the frozen operator and the 5th to 95th for differential expression. The operator's interval width is roughly two orders of magnitude narrower than that of differential expression.

| Estimator | Direction | Tissue | Median | 95% interval |
|---|---|---|---|---|
| Frozen operator | Inflammation | Colon | 0.9993 | 0.9989 to 0.9996 |
| Frozen operator | Inflammation | Ileum | 0.9990 | 0.9980 to 0.9993 |
| Frozen operator | IBD vs control | Colon | 0.9990 | 0.9977 to 0.9995 |
| Frozen operator | IBD vs control | Ileum | 0.9983 | 0.9967 to 0.9993 |
| Differential expression | Inflammation | Colon | 0.9813 | 0.9749 to 0.9886 |
| Differential expression | Inflammation | Ileum | 0.9512 | 0.8873 to 0.9679 |
| Differential expression | IBD vs control | Colon | 0.9537 | 0.8988 to 0.9655 |
| Differential expression | IBD vs control | Ileum | 0.8488 | 0.7122 to 0.9193 |

Supplementary Table 20 | Stability of the agreement between the frozen operator and differential expression, over 200 patient-blocked draws. The divergence on the IBD-state contrast is firm in colon and carries a wide interval in ileum.

| Direction | Tissue | Observed signed rho | Draw median | 90% interval | Draws at or below zero |
|---|---|---|---|---|---|
| Inflammation | Colon | 0.925 | 0.921 | 0.907 to 0.933 | 0 of 200 |
| Inflammation | Ileum | 0.735 | 0.728 | 0.671 to 0.775 | 0 of 200 |
| IBD vs control | Colon | 0.164 | 0.161 | 0.086 to 0.236 | 0 of 200 |
| IBD vs control | Ileum | 0.281 | 0.272 | 0.069 to 0.502 | 2 of 200 |

Supplementary Table 21 | GSE130038 pathway screen against time from diagnosis to colectomy, 26 patients. The complete 16-coordinate panel was screened, so the ranking rather than the nominal P value is the result. Remaining coordinates are in the source data.

| Frozen pathway coordinate | Spearman rho | Nominal P | Rank |
|---|---|---|---|
| Reactome mucin O-glycan biosynthesis | 0.486 | 0.012 | 1 of 16 |
| WNT and beta-catenin | 0.303 | | 2 of 16 |
| Independent full-transcriptome ssGSEA check | 0.507 | 0.0082 | |
| Panel genes present of 26 Reactome genes | 13 | | |

Supplementary Table 22 | GSE235236 cell-state signature provenance and alignment with frozen coordinates. Signatures were defined from the source study's published Source Data before any correlation with IBDex was inspected. The M2 null is what makes the other four interpretable.

| Signature | Source and rule | Strongest frozen coordinate | rho |
|---|---|---|---|
| IDA macrophage | top 10 cluster-specific genes, source study Fig. 2d data | TNF and NF-kappaB | 0.85 |
| Pan-neutrophil | four pan-neutrophil markers reported by the study | TNF and NF-kappaB | 0.82 |
| N3 interferon-neutrophil | top cluster-specific genes, source study Fig. 6c data | NOD1 and NOD2 | 0.79 |
| Inflammatory fibroblast | nine-gene panel displayed in source study Fig. 5d | NOD1 and NOD2 | 0.77 |
| M2 macrophage | top 10 cluster-specific genes, source study Fig. 2d data | none above 0.19 | 0.19 |

Supplementary Table 23 | Clinical remission strata against controls and against concordantly active disease. Scores are reduced to the patient median before testing. The disease-state displacement is invariant to the remission definition while the inflammation gap is not.

| Comparison | n | Axis | Median difference | AUC | P |
|---|---|---|---|---|---|
| Endoscopic remission vs control | 340 vs 241 | Inflammation | 0.404 | 0.655 | $1.7 \times 10^{-10}$ |
| Endoscopic remission vs control | 340 vs 241 | Disease state | 0.621 | 0.682 | $7.3 \times 10^{-14}$ |
| Histological remission vs control | 236 vs 241 | Inflammation | 0.586 | 0.716 | $3.3 \times 10^{-16}$ |
| Histological remission vs control | 236 vs 241 | Disease state | 0.629 | 0.702 | $2.3 \times 10^{-14}$ |
| Concordant remission vs control | 93 vs 241 | Inflammation | 0.120 | 0.581 | 0.021 |
| Concordant remission vs control | 93 vs 241 | Disease state | 0.668 | 0.725 | $1.8 \times 10^{-10}$ |
| Concordant remission vs concordantly active | 93 vs 252 | Inflammation | -1.485 | 0.165 | $1.2 \times 10^{-21}$ |
| Concordant remission vs concordantly active | 93 vs 252 | Disease state | 0.069 | 0.518 | 0.600 |

Supplementary Table 24 | Per-coordinate reliability of the frozen pathway atlas against independently computed single-sample enrichment. Reliability is declared for each coordinate separately because it is not uniform, and the two coordinates with insufficient panel coverage for an internal comparator are named rather than omitted.

| Frozen coordinate | Panel coverage | Colon rho | Ileum rho | Colon flag | Ileum flag |
|---|---|---|---|---|---|
| HALLMARK_INFLAMMATORY_RESPONSE | 115 of 200 | 0.694 | 0.767 | reliable | reliable |
| HALLMARK_TNFA_SIGNALING_VIA_NFKB | 110 of 200 | 0.733 | 0.604 | reliable | reliable |
| HALLMARK_IL6 | 50 of 87 | 0.613 | 0.719 | reliable | reliable |

| | | | | | |
|---|---|---|---|---|---|
| _JAK_STAT3_SIGNALING | | | | | |
| KEGG_CYTOKINE_CYTOKINE_RECEPTOR_INTERACTION | 123 of 295 | 0.674 | 0.750 | reliable | reliable |
| REACTOME_INTERLEUKIN_23_SIGNALING | 4 of 9 | not testable | not testable | - | - |
| REACTOME_NOD1_NOD2_SIGNALING_PATHWAY | 9 of 37 | 0.696 | 0.547 | reliable | reliable |
| REACTOME_EXTRACELLULAR_MATRIX_ORGANIZATION | 165 of 321 | 0.545 | 0.537 | reliable | reliable |
| HALLMARK_EPITHELIAL_MESENCHYMAL_TRANSITION | 131 of 200 | 0.640 | 0.619 | reliable | reliable |
| HALLMARK_TGF_BETA_SIGNALING | 11 of 54 | 0.525 | 0.520 | reliable | reliable |
| HALLMARK_PI3K_AKT_MTOR_SIGNALING | 16 of 105 | 0.574 | 0.415 | reliable | caution |
| HALLMARK_MTORC1_SIGNALING | 58 of 200 | 0.662 | 0.543 | reliable | reliable |
| HALLMARK_P53_PATHWAY | 54 of 200 | 0.446 | 0.150 | caution | unreliable |
| HALLMARK_WNT_BETA_CATENIN_SIGNALING | 8 of 42 | 0.647 | 0.706 | reliable | reliable |
| KEGG_BILE_SECRETION | 32 of 90 | 0.747 | 0.780 | reliable | reliable |
| HALLMARK_BILE_ACID_METABOLISM | 32 of 112 | 0.271 | 0.041 | unreliable | unreliable |
| REACTOME_TERMINATION_OF_O_GLYCAN_BIOSYNTHESIS | 13 of 26 | not testable | not testable | - | - |

Supplementary Table 25 | Independence of the sixteen frozen pathway coordinates across the reference. The atlas is not collinear overall, and the redundancy that exists is concentrated inside the inflammatory signaling family, which is therefore one axis carrying sub-readouts rather than six independent measurements.

| Compartment | Biopsies | Coordinate pairs | Median absolute rho | Pairs above 0.8 | Pairs above 0.6 | Strongest pair |
|---|---|---|---|---|---|---|
| Colon | 2,198 | 120 | 0.310 | 8 | 24 | Inflammatory response and IL6-JAK-STAT3, 0.99 |
| Ileum | 970 | 120 | 0.343 | 9 | 26 | Inflammatory response and cytokine-receptor interaction, 0.96 |

Supplementary Table 26 | Hallmark enrichment of the signed response rankings for both phenotype axes in both compartments. Weighted normalized enrichment scores are the values plotted in Fig. 3e; the unweighted variant is given alongside so the dissociation can be seen to be a property of the response maps rather than of the enrichment weighting. The false discovery rate is the smaller of the two compartments on the disease-state axis.

| Hallmark term | Inflammation colon | Inflammation ileum | Disease colon | Disease ileum | Inflammation colon, unweighted | Disease colon, unweighted | Disease FDR |
|---|---|---|---|---|---|---|---|
| Adipogenesis | -1.06 | 0.65 | 1.31 | 1.18 | -1.22 | 1.71 | 0.23 |
| Allograft Rejection | 1.98 | 2.03 | -1.17 | -1.39 | 3.91 | -2.15 | 0.0618 |
| Androgen | 1.33 | 0.95 | 1.37 | 1.20 | 1.03 | 1.05 | 0.202 |

| | | | | | | | |
|---|---|---|---|---|---|---|---|
| Response | | | | | | | |
| Angiogenesis | 1.66 | 1.56 | 0.55 | 0.93 | 1.87 | 1.13 | 0.846 |
| Apical Junction | 1.15 | 1.31 | 0.89 | -0.69 | 1.26 | 0.90 | 0.868 |
| Apical Surface | 1.10 | 0.94 | 1.00 | 0.83 | 1.00 | 0.77 | 0.623 |
| Apoptosis | 1.50 | 1.35 | 1.61 | 1.50 | 1.29 | 1.21 | 0.0202 |
| Bile Acid Metabolism | -1.02 | -1.36 | 0.61 | 0.87 | -1.60 | 1.21 | 0.851 |
| Cholesterol Homeostasis | 1.55 | 1.13 | 2.06 | 1.84 | 1.67 | 2.05 | 0.00331 |
| Coagulation | 1.63 | 1.47 | 0.82 | 0.87 | 1.70 | 1.74 | 0.851 |
| Complement | 1.92 | 1.93 | 0.94 | 0.82 | 2.09 | 0.59 | 0.73 |
| E2F Targets | 1.42 | 1.37 | 3.03 | 3.18 | 1.99 | 5.13 | 0.00331 |
| Epithelial Mesenchymal Transition | 1.64 | 1.57 | -0.47 | -0.57 | 2.28 | 1.63 | 1 |
| Estrogen Response Early | 0.86 | 1.06 | 1.05 | 1.38 | 0.64 | 1.20 | 0.0626 |
| Estrogen Response Late | 1.01 | 1.01 | 1.71 | 1.74 | -1.09 | 2.04 | 0.00497 |
| Fatty Acid Metabolism | 0.97 | -0.99 | 1.89 | 1.85 | -1.29 | 2.47 | 0.00331 |
| G2M Checkpoint | 1.49 | 1.59 | 3.12 | 3.16 | 2.56 | 5.04 | 0.00331 |
| Glycolysis | 1.24 | 0.66 | 1.80 | 1.51 | 0.96 | 2.50 | 0.00331 |
| Hedgehog Signaling | -0.95 | -0.77 | -1.13 | -0.85 | -0.72 | -0.70 | 0.494 |
| Heme Metabolism | 0.85 | 1.09 | 1.02 | 0.85 | -0.80 | 1.24 | 0.614 |
| Hypoxia | 1.08 | 0.80 | 0.79 | 0.88 | 0.81 | 1.01 | 0.851 |
| IFN-alpha | 2.18 | 2.34 | 2.60 | 2.40 | 3.31 | 4.38 | 0.00331 |
| IFN-gamma | 2.47 | 2.53 | 2.04 | 1.70 | 4.38 | 2.67 | 0.00331 |
| IL2-STAT5 | 1.77 | 2.00 | -1.42 | -1.49 | 2.18 | -2.01 | 0.0449 |
| IL6-JAK-STAT3 | 2.03 | 2.00 | 0.68 | 0.69 | 2.45 | 0.91 | 0.994 |
| Inflammatory | 2.18 | 2.24 | -1.05 | -1.01 | 3.83 | -1.32 | 0.546 |
| KRAS up | 1.24 | 1.53 | -1.09 | -0.96 | 1.22 | -1.35 | 0.481 |
| Kras Signaling Dn | -1.23 | -1.49 | -0.79 | -0.84 | -1.99 | 0.68 | 0.851 |
| MTORC1 | 2.18 | 1.85 | 1.84 | 1.56 | 3.26 | 2.02 | 0.00331 |
| Mitotic Spindle | 1.51 | 1.43 | 2.60 | 2.30 | 2.12 | 3.06 | 0.00331 |
| Myc Targets V1 | 1.24 | 1.47 | 2.15 | 2.20 | 1.54 | 2.12 | 0.00331 |
| Myogenesis | 0.65 | -0.90 | 1.54 | 1.43 | -1.63 | 1.69 | 0.0335 |
| P53 Pathway | 1.67 | 1.55 | 1.43 | 1.51 | 1.24 | 1.09 | 0.0449 |
| Pancreas Beta Cells | -1.68 | -1.82 | -0.70 | 1.49 | -1.78 | -1.26 | 0.124 |
| Peroxisome | -0.85 | -0.97 | 1.29 | 1.30 | -1.10 | 1.66 | 0.238 |
| Pi3K Akt Mtor Signaling | 1.31 | 1.30 | 1.02 | 1.18 | 1.06 | -0.91 | 0.432 |
| Reactive Oxygen Species Pathway | 1.40 | 1.18 | 1.14 | 0.87 | 1.66 | 1.01 | 0.494 |
| Spermatogenesis | 0.59 | 0.61 | 2.01 | 2.05 | 0.57 | 1.61 | 0.00331 |
| TNF/NF-kB | 2.33 | 2.46 | -1.59 | -0.87 | 3.84 | -2.30 | 0.00497 |
| Tgf Beta Signaling | 0.87 | 0.78 | -0.68 | 0.90 | 0.87 | -0.61 | 0.851 |
| UPR | 2.07 | 1.97 | 1.41 | 2.00 | 2.70 | 1.25 | 0.00954 |
| Uv Response Dn | 1.26 | 0.94 | -0.79 | -0.52 | 1.42 | -1.11 | 0.974 |
| Uv Response Up | 0.98 | 1.08 | 1.49 | 1.52 | 0.72 | 1.22 | 0.0496 |
| Xenobiotic | -1.12 | -1.53 | 1.83 | 1.73 | -2.22 | 2.14 | 0.00331 |

#### Supplementary Figures

Figure S1. Study selection and corpus assembly. PRISMA-style flow from records identified across the Gene Expression Omnibus, the IBD Multi'omics Database and the IBDome consortium, through sample-level eligibility assessment, to the four construction cohorts. Per-cohort composition by diagnosis, anatomical site and inflammatory annotation.

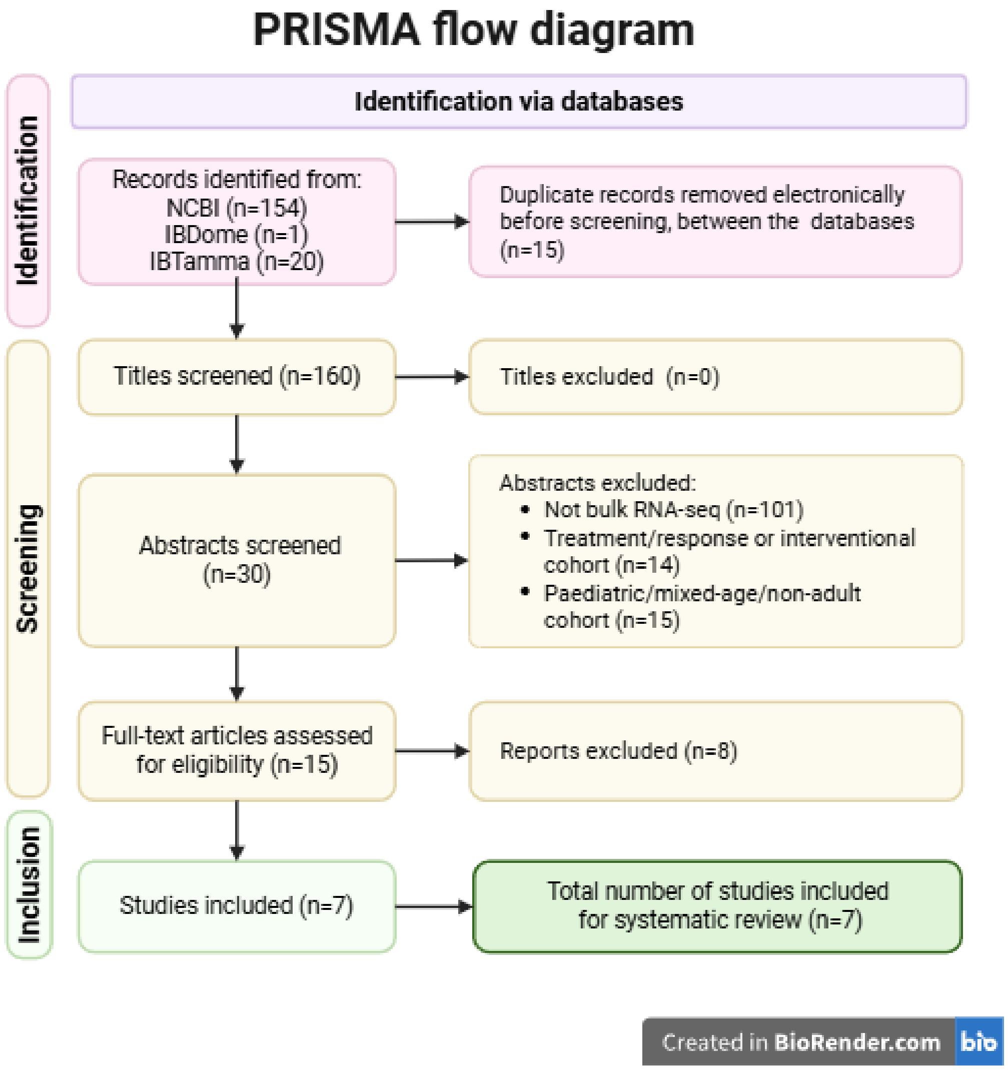


Figure S2. Linear baselines remain organized by cohort. PCA and UMAP of the standardized 3,000-gene interface colored by dataset, anatomy and diagnosis, repeated after ComBat-seq-style per-dataset adjustment, against the frozen latent space colored identically. Cohort and compartment dominate the linear views throughout while diagnoses overlap, which is the observation that motivates suppressing provenance during training rather than as a preprocessing step.

**a** **b** **c**

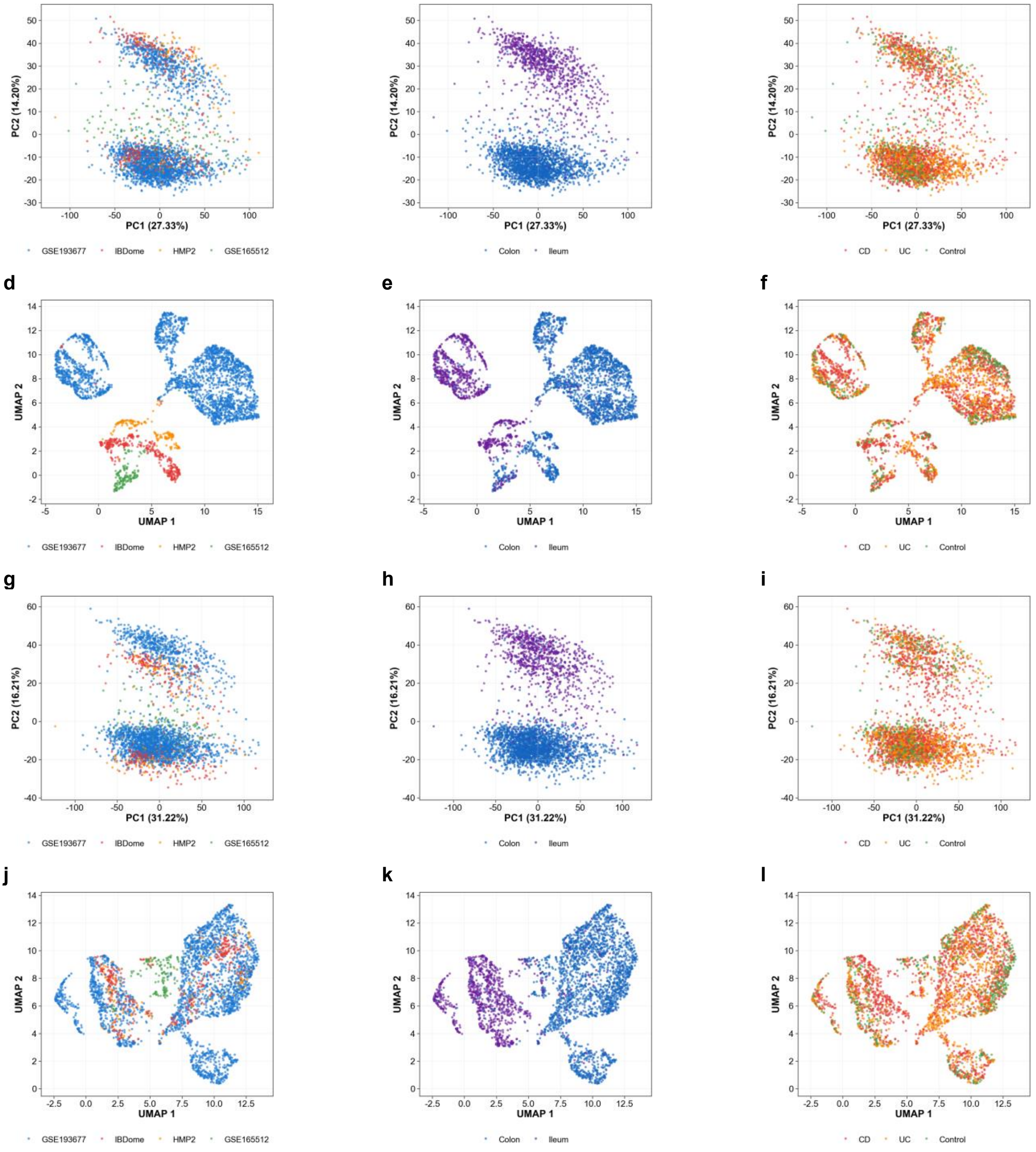


Figure S3. Training diagnostics and the per-dimension provenance audit. a, Training and validation objective across epochs, with the selected checkpoint at epoch 179 and early stopping at 209 marked. b, Validation objective decomposed into reconstruction, Kullback-Leibler and the three adversarial terms, showing the annealing schedules. c, Partial R-squared of dataset, diagnosis and anatomical site against each of the 16 latent dimensions. d, Variance carried by each dimension.

**a**

**b**

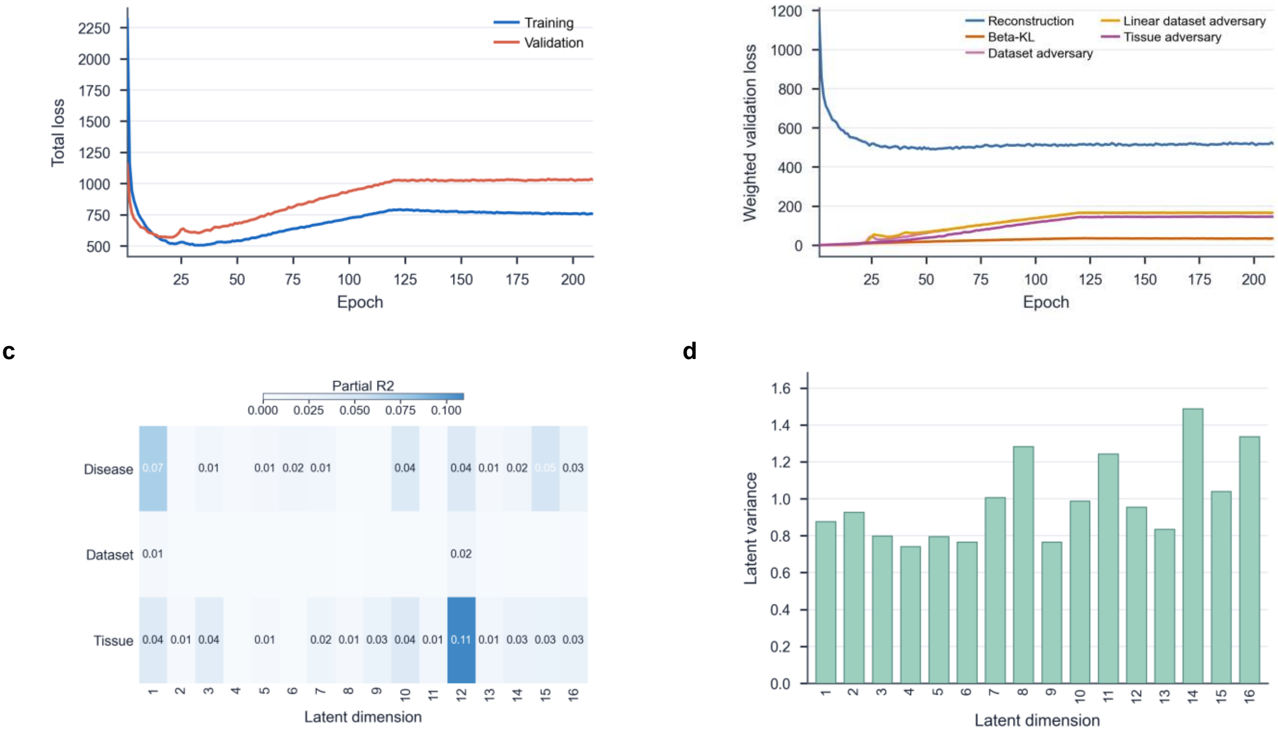


Figure S4. Consensus regions and their composition. a, Colon regions on the frozen latent embedding. b,c, Composition of the colon regions by cohort of origin and by diagnosis. d, Ileal regions. e,f, Composition of the ileal regions. Every region draws from more than one cohort and contains all three diagnostic categories.

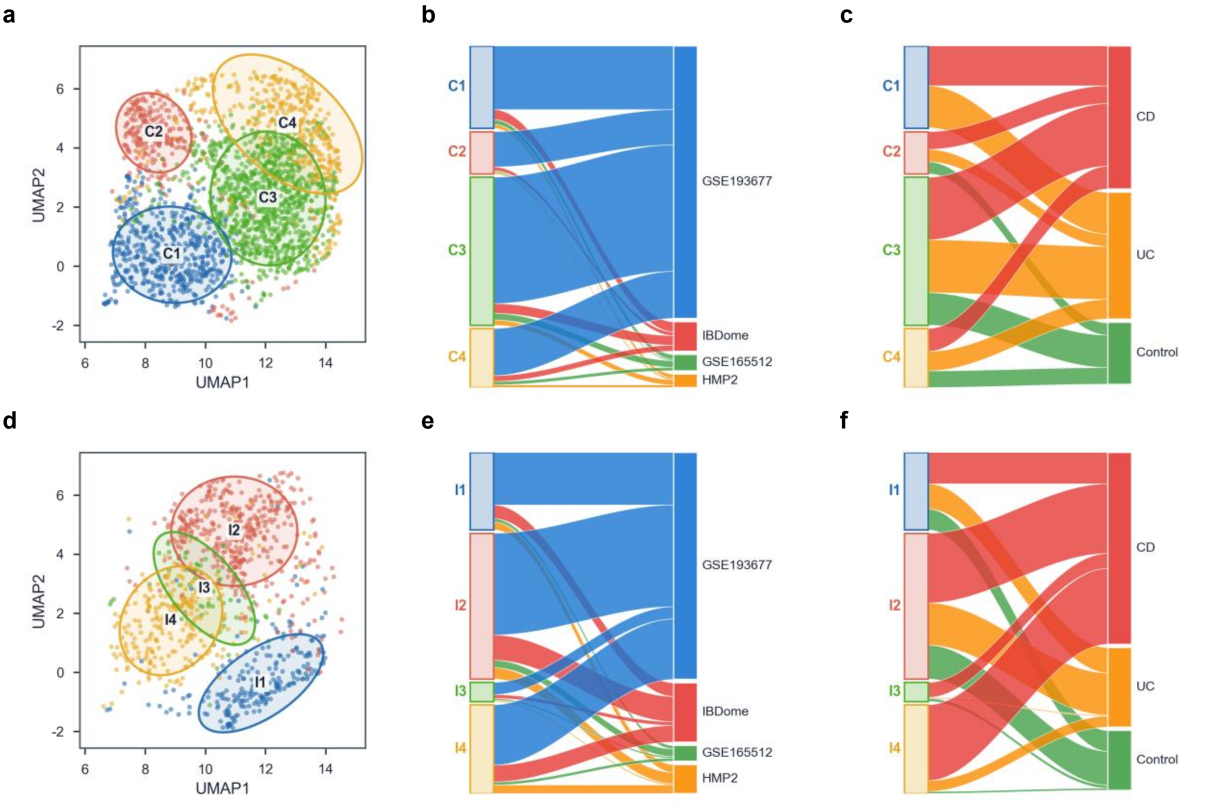

Figure S5. The sixteen frozen pathway coordinates against independently computed ssGSEA. Spearman correlation across all 3,168 reference biopsies, shown separately for colon and ileum, with panel coverage given at the right as the number of gene-set members present in the frozen 3,000-gene interface. Bands mark the reliability call carried in Supplementary Table 24. Twelve of the fourteen testable coordinates are reliable in colon and eleven in ileum, where Hallmark PI3K-AKT-mTOR signaling is flagged caution. Hallmark p53 pathway and Hallmark bile acid metabolism are declared weak rather than quietly reported, and the two coordinates whose gene sets are too sparsely covered to test are marked as such. Only the sixteen coordinates released with the reference are shown.

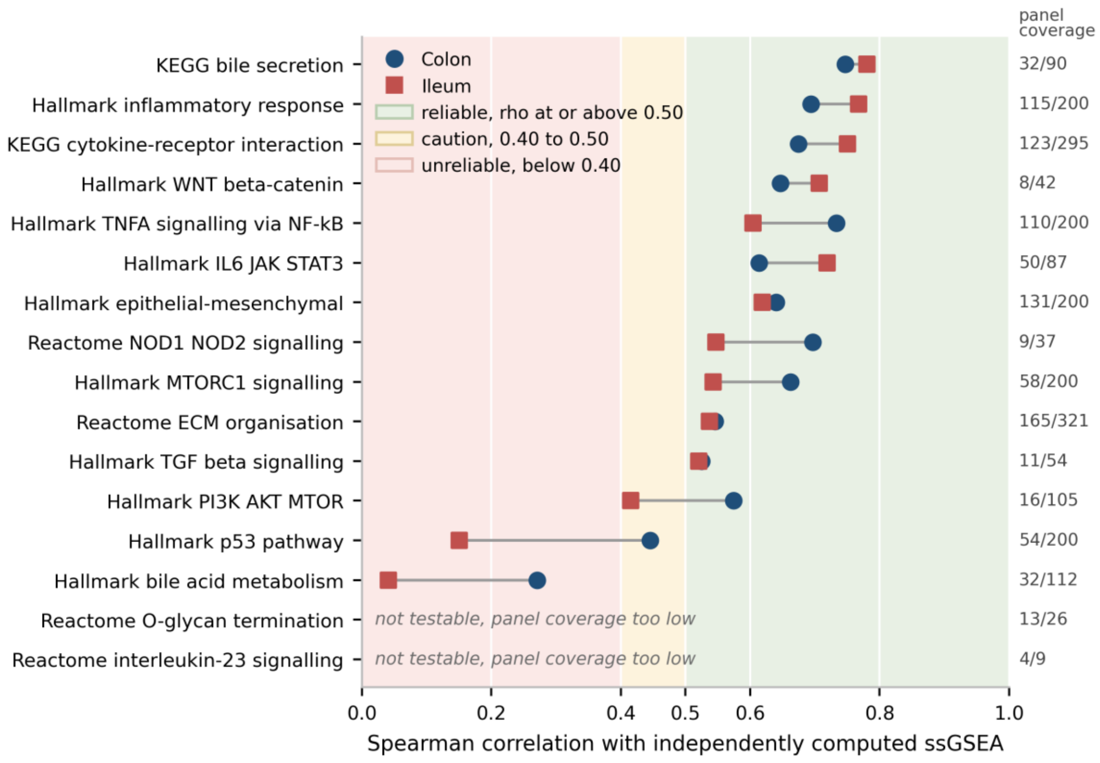


Figure S6. The response operator is a property of decoder geometry rather than of how it was estimated. All correlations are signed. a,b, Signed colon-versus-ileum rank correlation of the inflammation response map under finite-difference step sizes spanning two orders of magnitude, and under three rules for combining site-level Jacobians into compartment operators. c,d, Signed rank correlation between the reported map and the map recovered under each alternative, which is the quantity that decides whether the choice matters. Step size leaves the ranking identical to four decimal places. Equal site weighting and elementwise median aggregation move it by 0.0044 and 0.0059, so the compartment operator is not an artifact of the site that contributed most samples. Filled markers are the reported settings. Per-gene bootstrap stability over 200 patient-level draws within tissue is reported separately.

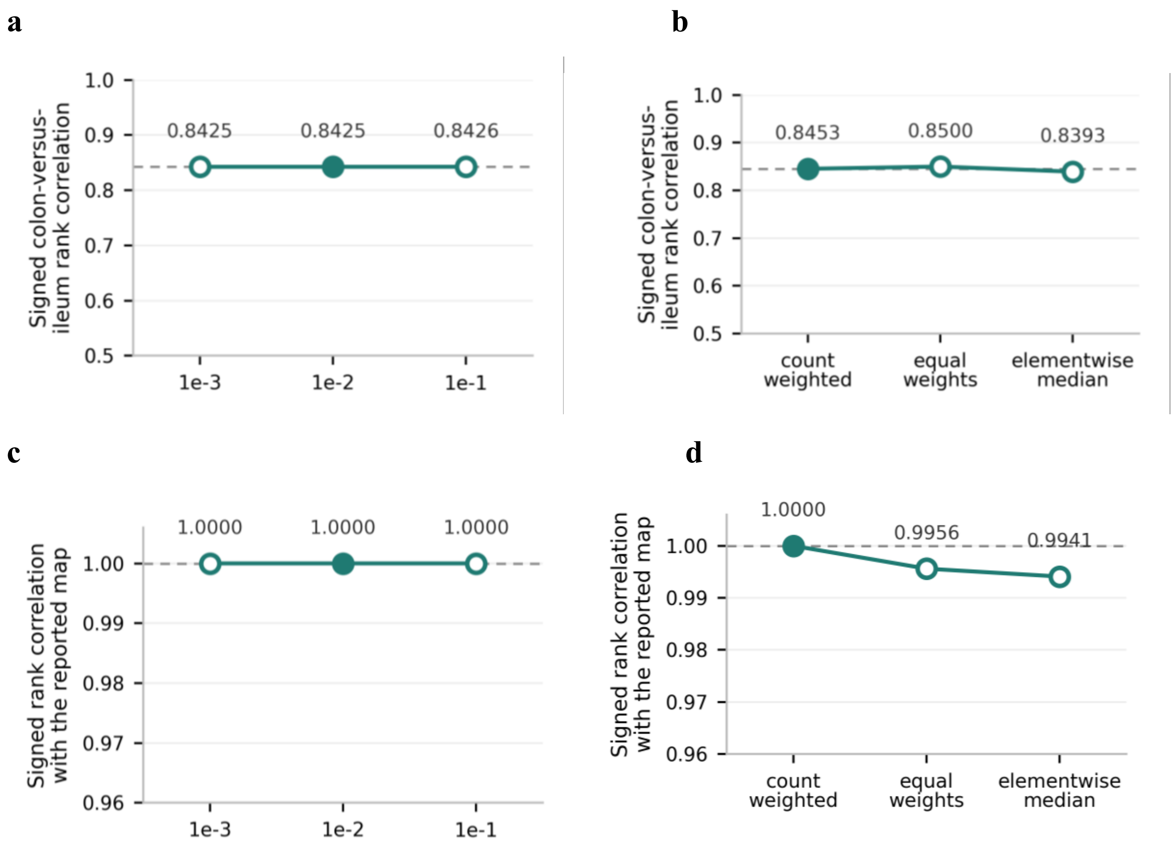
a
Signed colon-versus-ileum rank correlation
1.0
0.9
0.8
0.7
0.6
0.5
0.8425
0.8425
0.8426
1e-3
1e-2
1e-1
b
Signed colon-versus-ileum rank correlation
0.8453
0.8500
0.8393
count weighted
equal weights
elementwise median
c
Signed rank correlation with the reported map
1.00
0.99
0.98
0.97
0.96
1.0000
1.0000
1.0000
1e-3
1e-2
1e-1
d
Signed rank correlation with the reported map
1.0000
0.9956
0.9941
count weighted
equal weights
elementwise median
